# Gastrointestinal medication use patterns and incident depressive symptoms among adults aged 50 years or older: a prospective multicohort study

**DOI:** 10.64898/2026.10.26.26366451

**Authors:** Shupeng Wang, Dongdong Lu, Tao Tao

## Abstract

**Background:** Depressive symptoms contribute substantially to disability and loss of independence in later life, and gastrointestinal illness frequently co-occurs with affective vulnerability. However, evidence linking gastrointestinal medication use to subsequent depressive symptoms remains largely confined to single countries and cross-sectional assessments. We investigated whether baseline gastrointestinal medication use and longitudinal two-wave medication-use patterns are associated with incident depressive symptoms across three prospective ageing cohorts.

**Methods:** We analysed community-dwelling adults aged 50 years or older from the China Health and Retirement Longitudinal Study (CHARLS), the US Health and Retirement Study (HRS), and the Survey of Health, Ageing and Retirement in Europe (SHARE) who scored below cohort-specific depressive-symptom thresholds at baseline, followed for a median of 8–11 years. Baseline medication use was compared with non-use; two-wave patterns comprised continued non-use (reference), initiation, discontinuation, and persistent use at both waves. Cohort-specific Cox proportional hazards models with cluster-robust standard errors adjusted for sociodemographic, behavioural, and chronic health covariates (multiple imputation for missing data); estimates were synthesised using random-effects meta-analysis. Negative- and positive-control medications and Fine–Gray competing-risk regression models evaluated specificity and survival bias.

**Results:** The baseline analysis included 50,467 participants (median age 59–65 years; 44.4%–56.8% female) with 16,896 incident depressive events; the two-wave analysis included 32,686 participants with 8,069 events. Pooled adjusted hazard ratios (HRs) were 1.31 (95% CI 1.21–1.41; *I*² = 29%) for baseline use and 1.34 (95% CI 1.22–1.47; *I*² = 0%) for persistent use; initiation (1.27, 1.13–1.43) and discontinuation (1.25, 1.11–1.40) showed consistent risk elevations across cohorts. Antihypertensive and lipid-lowering medications (negative controls) showed null associations with incident depression, whereas psychiatric medications (positive control) showed the expected strong relationship (pooled HR 1.99, 95% CI 1.79–2.22). The association remained robust after further adjusting for continuous baseline subthreshold depressive symptoms (pooled HR 1.21, 95% CI 1.14–1.27; *I*² = 0%), and medicated digestive disease carried higher risk than untreated digestive disease (pooled HR 1.18, 95% CI 1.05–1.32; *I*² = 0%). Accounting for competing mortality did not alter the estimates.

**Conclusions:** Baseline and persistent gastrointestinal medication use were consistently associated with increased risk of incident depressive symptoms across three cross-national ageing cohorts with negligible heterogeneity. These findings support integrating routine affective screening into medication reviews for older adults receiving gastrointestinal pharmacotherapy, while maintaining guideline-directed management of digestive conditions.

## Background

Depressive symptoms are a leading cause of disability, healthcare utilisation, and loss of independence in later life, making their prevention a public health priority in ageing populations [1, 2]. Concurrently, gastrointestinal complaints—including gastro-oesophageal reflux, dyspepsia, abdominal pain, and altered bowel habits—are among the most frequent reasons for primary care consultation in older adults and commonly co-occur with anxiety and depression [3–6]. Given the bidirectional interplay between digestive morbidity and affective regulation, the clinical management of gastrointestinal symptoms represents a critical window of psychological vulnerability [7].

Gastrointestinal medications are prescribed on a global scale, with proton pump inhibitors ranking among the most widely used drug classes worldwide—often without a documented clinical indication [8]. Biological pathways potentially linking long-term acid suppression to affective disturbance include drug-induced perturbations in gut microbiota composition and short-chain fatty acid production, altered gut–brain axis signalling, and impaired absorption of micronutrients such as vitamin B12 and magnesium [9, 10]. Supporting this hypothesis, observational studies have identified a higher prevalence of depression among users of prescription medications with potential depressogenic effects, including proton pump inhibitors [11], and population-based surveys have linked gastric acid suppression to depressive symptoms and suicidal ideation [12, 13]. Registry and cohort studies have extended these findings to perinatal populations and active-comparator analyses of acid-suppressing agents [14, 15], alongside meta-analytic and genetic evidence demonstrating bidirectional links between gastro-oesophageal reflux and affective disorders [16, 17]. Conversely, other longitudinal observations suggest that depressive-symptom trajectories may track the severity of underlying digestive morbidity and its symptomatic relief during therapy rather than a direct pharmacological effect [18–20].

Three critical research gaps warrant investigation. First, most prior studies have assessed medication exposure cross-sectionally at a single timepoint, precluding differentiation among treatment initiation, discontinuation, and persistent use—longitudinal patterns that capture distinct clinical courses. Second, existing evidence is predominantly restricted to single-country studies, leaving it unclear whether associations reflect regional prescribing practices or a generalisable phenomenon. Third, medication reports in population surveys may reflect underlying morbidity and healthcare utilisation; yet, few studies have evaluated specificity against active negative-control medications or distinguished pharmacotherapy from underlying disease. Addressing these gaps is critical in older populations, where polypharmacy and digestive complaints frequently co-occur and depressive symptoms are often under-recognised.

Therefore, leveraging harmonised longitudinal data from three national ageing cohorts—the China Health and Retirement Longitudinal Study (CHARLS) in China, the Health and Retirement Study (HRS) in the United States, and the Survey of Health, Ageing and Retirement in Europe (SHARE) across 16 European countries [21–23]—we aimed to: (1) delineate the associations of baseline gastrointestinal medication use and two-wave medication-use patterns (initiation, discontinuation, and persistent use at both waves) with incident depressive symptoms and quantify their cross-cohort consistency; (2) evaluate the specificity of these associations by stratifying by digestive-disease diagnosis and testing four negative-control medication classes alongside a positive-control exposure; and (3) assess the robustness of the findings after accounting for baseline subthreshold depressive symptoms and all-cause mortality as a competing risk. We hypothesised that baseline and persistent two-wave gastrointestinal medication use would be associated with a higher incidence of depressive symptoms, independently of underlying digestive morbidity and healthcare-contact confounding.

## Methods

### Study design and participants

We conducted parallel prospective cohort analyses using harmonised longitudinal data from CHARLS [21], HRS [22], and SHARE [23]. Baseline assessments (t0) corresponded to CHARLS wave 1 (2011), HRS wave 11 (2012), and SHARE wave 4 (2010/11), with second-round medication assessments (t1) conducted at CHARLS wave 2 (2013), HRS wave 12 (2014), and SHARE wave 5 (2013). Depressive symptoms were tracked through the 2020 CHARLS survey, HRS wave 15 (2020), and SHARE wave 9 (2021/22); SHARELIFE (wave 7, 2016/17) collected retrospective life-history data only and was excluded from outcome ascertainment. To account for differences in questionnaire wording, depressive-symptom scales, survey schedules, and healthcare systems, cohorts were analysed separately prior to meta-analytic synthesis (Table S1).

Two prospective analytic cohorts were constructed (Fig. 1 and Fig. S1). For the baseline analysis, eligible participants were aged 50 years or older at t0, had complete baseline medication and depressive-symptom data, scored below the cohort-specific depressive-symptom threshold at t0, and completed at least one follow-up depressive-symptom assessment (6,452 in CHARLS, 12,938 in HRS, and 31,077 in SHARE; total *n* = 50,467). The two-wave analysis additionally required participation at t1, valid medication reports at both t0 and t1, depressive-symptom scores below threshold at both waves, and at least one subsequent symptom assessment after t1 (4,087 in CHARLS, 9,801 in HRS, and 18,798 in SHARE; total *n* = 32,686). Up to four follow-up waves were available in CHARLS and HRS and up to five in SHARE. SHARE contributed 16 European countries to the baseline analysis and 13 countries with repeated medication assessments to the two-wave analysis (Table S21). This study followed the Strengthening the Reporting of Observational Studies in Epidemiology (STROBE) guideline (Additional file 2).

**Figure 1.**
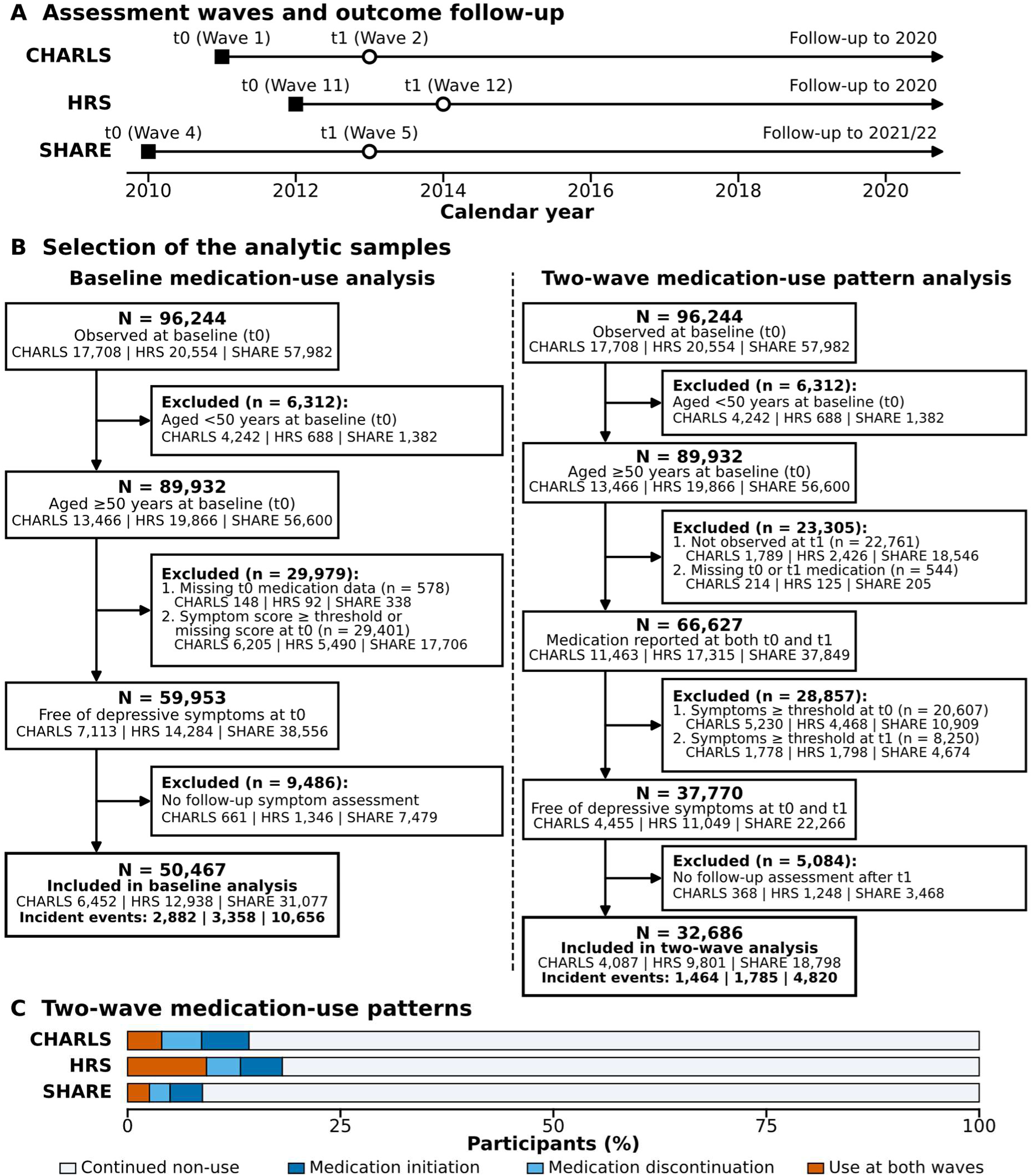
Study design and analytic samples. (A) Assessment waves and outcome follow-up timelines in CHARLS, HRS and SHARE; solid squares denote the baseline medication assessment (t0), open circles denote the second-round medication assessment (t1), and horizontal arrows indicate the outcome follow-up period. (B) Selection of the baseline (*n* = 50,467) and two-wave (*n* = 32,686) analytic samples; each box gives the number remaining (total, with the cohort-specific CHARLS, HRS and SHARE values shown beneath), and each exclusion card shows the number excluded at that step, split by cohort. The final exclusion step reflects absence of a valid follow-up depressive-symptom assessment (loss to follow-up or death before the first scheduled interview). Cohort-specific flows are shown in Fig. S1. (C) Proportions of the four two-wave gastrointestinal medication-use patterns in each cohort (exact counts and percentages are provided in Table 1, panel B).

**Table 1.** Baseline characteristics of participants in the baseline and two-wave gastrointestinal medication-use analyses, by cohort.

**A. Baseline medication-use analysis**
| Characteristic | CHARLS (*n*=6,452) | HRS (*n*=12,938) | SHARE (*n*=31,077) |
| --- | --- | --- | --- |
| Age, years, median (IQR) | 59 (55–66) | 65 (58–74) | 64 (58–71) |
| Female sex, *n* (%) | 2,862 (44.4%) | 7,352 (56.8%) | 16,145 (52.0%) |
| Gastrointestinal medication use, *n* (%) | 577 (8.9%) | 1,849 (14.3%) | 1,680 (5.4%) |
| **Education, *n* (%)** |  |  |  |
| Low | 5,552 (86.1%) | 1,892 (14.6%) | 11,553 (37.2%) |
| Middle | 755 (11.7%) | 7,700 (59.5%) | 12,423 (40.0%) |
| High | 145 (2.2%) | 3,343 (25.8%) | 7,101 (22.8%) |
| Currently employed, *n* (%) | 3,935 (61.8%) | 5,793 (44.8%) | 11,258 (36.4%) |
| **Household wealth tertile, *n* (%)** |  |  |  |
| Low | 1,512 (32.8%) | 4,286 (33.1%) | 9,752 (31.4%) |
| Middle | 1,574 (34.2%) | 4,316 (33.4%) | 10,469 (33.7%) |
| High | 1,522 (33.0%) | 4,336 (33.5%) | 10,856 (34.9%) |
| **Household and family structure, *n* (%)** |  |  |  |
| Married/partnered | 5,771 (89.4%) | 8,137 (62.9%) | 22,258 (72.7%) |
| Co-residing with at least one child | 3,532 (56.0%) | 3,542 (27.4%) | 6,314 (22.3%) |
| **Smoking status, *n* (%)** |  |  |  |
| Never | 3,615 (56.1%) | 5,888 (45.7%) | 16,015 (52.3%) |
| Former | 649 (10.1%) | 5,401 (42.0%) | 8,930 (29.2%) |
| Current | 2,185 (33.9%) | 1,583 (12.3%) | 5,660 (18.5%) |
| Current/recent alcohol use, *n* (%) | 2,339 (36.3%) | 7,557 (58.4%) | 19,493 (63.0%) |
| BMI, kg/m<sup>2</sup>, mean (SD) | 23.6 (3.8) | 28.5 (5.8) | 26.8 (4.4) |
| Depressive-symptom score at risk origin, mean (SD) | 4.41 (2.78) | 0.56 (0.72) | 1.35 (1.07) |
| **Chronic-condition burden, *n* (%)** |  |  |  |
| 0 | 2,845 (44.9%) | 2,232 (17.3%) | 11,127 (35.8%) |
| ≥1 | 3,498 (55.1%) | 10,706 (82.7%) | 19,944 (64.2%) |
| Self-reported digestive disease, *n* (%) | 1,080 (16.7%) | NA | 1,796 (5.8%) |
| Person-years of follow-up | 39,725 | 75,700 | 209,762 |
| Median follow-up, years | 9 | 8 | 11 |
| Incident depressive symptoms, *n* (%) | 2,882 (44.7%) | 3,358 (26.0%) | 10,656 (34.3%) |
| Incidence rate, per 1,000 person-years | 72.6 | 44.4 | 50.8 |

B. Two-wave medication-use pattern analysis
| Characteristic | CHARLS (*n*=4,087) | HRS (*n*=9,801) | SHARE (*n*=18,798) |
| --- | --- | --- | --- |
| Age, years, median (IQR) | 59 (55–65) | 65 (58–73) | 64 (58–71) |
| Female sex, *n* (%) | 1,693 (41.4%) | 5,490 (56.0%) | 9,465 (50.4%) |
| **Two-wave medication-use pattern, *n* (%)** |  |  |  |
| Continued non-use | 3,504 (85.7%) | 8,019 (81.8%) | 17,142 (91.2%) |
| Medication initiation | 227 (5.6%) | 482 (4.9%) | 718 (3.8%) |
| Medication discontinuation | 192 (4.7%) | 390 (4.0%) | 456 (2.4%) |
| Use reported at both waves | 164 (4.0%) | 910 (9.3%) | 482 (2.6%) |
| **Education, *n* (%)** |  |  |  |
| Low | 3,492 (85.4%) | 1,259 (12.8%) | 6,326 (33.7%) |
| Middle | 506 (12.4%) | 5,818 (59.4%) | 7,654 (40.7%) |
| High | 89 (2.2%) | 2,721 (27.8%) | 4,818 (25.6%) |
| Currently employed, *n* (%) | 2,602 (64.4%) | 4,685 (47.8%) | 7,389 (39.4%) |
| **Household wealth tertile, *n* (%)** |  |  |  |
| Low | 966 (32.8%) | 3,266 (33.3%) | 6,154 (32.7%) |
| Middle | 982 (33.4%) | 3,229 (32.9%) | 6,219 (33.1%) |
| High | 995 (33.8%) | 3,306 (33.7%) | 6,425 (34.2%) |
| **Household and family structure, *n* (%)** |  |  |  |
| Married/partnered | 3,723 (91.1%) | 6,372 (65.0%) | 13,527 (72.9%) |
| Co-residing with at least one child | 2,211 (55.2%) | 2,721 (27.8%) | 3,563 (20.8%) |
| **Smoking status, *n* (%)** |  |  |  |
| Never | 2,227 (54.5%) | 4,540 (46.5%) | 9,694 (52.2%) |
| Former | 427 (10.5%) | 4,124 (42.3%) | 5,574 (30.0%) |
| Current | 1,431 (35.0%) | 1,090 (11.2%) | 3,287 (17.7%) |
| Current/recent alcohol use, *n* (%) | 1,574 (38.5%) | 5,932 (60.5%) | 12,582 (67.1%) |
| BMI, kg/m<sup>2</sup>, mean (SD) | 23.7 (3.7) | 28.6 (5.7) | 26.7 (4.3) |
| Depressive-symptom score at risk origin, mean (SD) | 4.16 (2.76) | 0.49 (0.68) | 1.22 (1.05) |
| **Chronic-condition burden, *n* (%)** |  |  |  |
| 0 | 1,901 (47.2%) | 1,842 (18.8%) | 7,165 (38.1%) |
| ≥1 | 2,124 (52.8%) | 7,959 (81.2%) | 11,631 (61.9%) |
| Self-reported digestive disease, *n* (%) | 667 (16.3%) | NA | 999 (5.3%) |
| Person-years of follow-up | 22,001 | 48,110 | 120,507 |
| Median follow-up, years | 7 | 6 | 9 |
| Incident depressive symptoms, *n* (%) | 1,464 (35.8%) | 1,785 (18.2%) | 4,820 (25.6%) |
| Incidence rate, per 1,000 person-years | 66.5 | 37.1 | 40.0 |

Percentages were calculated using available non-missing denominators, which varied by characteristic. Household wealth tertiles were defined within each cohort-analysis eligible sample. Chronic-condition burden comprised hypertension, diabetes, cancer, chronic lung disease, heart disease, stroke, mental illness and arthritis. BMI, body mass index; CHARLS, China Health and Retirement Longitudinal Study; HRS, Health and Retirement Study; IQR, interquartile range; SHARE, Survey of Health, Ageing and Retirement in Europe. Depressive-symptom scores were measured with the 10-item Center for Epidemiologic Studies Depression scale (CESD-10, range 0–30) in CHARLS, the 8-item CESD (CESD-8, range 0–8) in HRS and the EURO-D scale (range 0–12) in SHARE; therefore, scores are not directly comparable across cohorts. A harmonised self-reported digestive-disease diagnosis was not available in HRS and is shown as NA.

### Medication exposure

The primary exposure was self-reported current use of medication for stomach or other digestive conditions (hereafter gastrointestinal medication use), operationalised using the corresponding survey item in each cohort (Table S1). In CHARLS, participants reporting a physician-diagnosed stomach or digestive disease indicated whether they were currently taking Western modern medication for the condition; a prespecified sensitivity analysis additionally incorporated traditional Chinese medicine and over-the-counter digestive treatments (Table S1). In HRS, exposure was ascertained from the harmonised stomach and gastrointestinal medication indicator derived from regular prescription medication reports. In SHARE, participants reported whether they currently took medications at least once weekly for stomach burns or ulcers. Cross-classification of baseline digestive-disease diagnosis and medication status in CHARLS and SHARE is detailed in Table S18.

Exposure was evaluated under two complementary frameworks. The baseline analysis compared current gastrointestinal medication users with non-users at t0. The two-wave longitudinal analysis classified medication status across t0 and t1 into four mutually exclusive patterns: continued non-use (reference), medication initiation (non-use at t0 and use at t1), medication discontinuation (use at t0 and non-use at t1), and persistent use (use reported at both t0 and t1) (Fig. 1).

### Depressive-symptom outcome and follow-up

Incident depressive symptoms were defined as the first follow-up survey at which a participant’s depressive-symptom score reached or exceeded the validated cohort-specific cut-off. Depressive symptoms were assessed using the 10-item Center for Epidemiologic Studies Depression Scale (CES-D-10; range 0–30, threshold ≥10) in CHARLS, the 8-item CES-D (CES-D-8; range 0–8, threshold ≥3) in HRS, and the 12-item EURO-D scale (range 0–12, threshold ≥4) in SHARE [24, 25]. All participants scored below the respective threshold at the risk origin (t0 for the baseline analysis; both t0 and t1 for the two-wave analysis).

Time at risk commenced at t0 for the baseline analysis and at t1—following completion of both exposure-assessment waves—for the two-wave analysis. Follow-up continued until the first incident depressive-symptom event or the final survey wave with a complete symptom assessment, whichever occurred first; participants without an event were right-censored at their last valid assessment. To account for interval censoring across biennial or triennial survey waves, we conducted sensitivity analyses using discrete-time complementary log–log regression models fitted to the observed survey intervals.

## Covariates

Covariates were ascertained at baseline (t0) in both analytical frameworks to prevent overadjustment for intermediate variables affected by initial medication use.

Sociodemographic covariates comprised age (continuous, in years), sex (male or female), harmonised educational attainment (low, middle, or high), current paid employment (yes or no), within-cohort household wealth tertile (low, middle, or high), marital or partnership status (partnered vs unpartnered), and co-residence with any child (yes or no). Lifestyle and anthropometric factors included smoking status (never, former, or current), alcohol consumption (current/recent drinker vs non-drinker), and continuous body mass index (BMI, kg/m²).

Baseline chronic-condition burden was derived from eight physician-diagnosed conditions— hypertension, diabetes, cancer, chronic lung disease, heart disease, stroke, psychiatric illness, and arthritis—and dichotomised as 0 versus ≥1 condition (reference: 0). Four nested adjustment models were prespecified: Model 1 was unadjusted; Model 2 adjusted for age and sex; Model 3 additionally adjusted for educational attainment, employment status, household wealth tertile, partnership status, and co-residence with children; and Model 4 (primary model) further adjusted for smoking status, alcohol consumption, BMI, and chronic-condition burden.

### Missing data

Missing covariate data were imputed separately within each cohort-specific analytical subset using multiple imputation by chained equations (MICE). Conditional imputation models were tailored to each variable’s measurement scale: logistic regression for binary predictors, multinomial logistic regression for nominal unordered categorical variables, proportional-odds ordinal regression for ordered categorical variables, and predictive mean matching for continuous variables such as BMI. Each imputation model incorporated all covariates from the primary outcome regression model, alongside the exposure variable, incident event status, count of completed follow-up surveys applied for participant eligibility, and matched covariate data captured from the preceding survey wave [26, 27]. Analyses were performed using R version 4.5.3 with the survival, mice, metafor and cmprsk packages. We generated 20 imputed datasets and combined model estimates using Rubin’s rules.

### Statistical analysis

Baseline characteristics were summarised separately for the baseline and two-wave analytic samples as means (standard deviations [SDs]) or medians (interquartile ranges [IQRs]) for continuous variables and as counts (percentages) among non-missing observations for categorical variables. Exposure-stratified baseline characteristics are presented in Tables S2–S7.

### Primary Cox proportional-hazards analyses

Cohort-specific Cox proportional-hazards regression models were fitted to estimate hazard ratios (HRs) and 95% confidence intervals (CIs) for incident depressive symptoms. Follow-up time in years from the risk origin (t0 for baseline analyses and t1 for two-wave analyses) served as the underlying timescale, with cluster-robust standard errors accounting for within-household clustering. In SHARE, country was included as a fixed effect to estimate within-country associations. The primary fully adjusted model (Model 4) controlled for age, sex, education, employment, household wealth, partnership status, co-residence with children, smoking status, alcohol consumption, BMI, and binary chronic-condition burden (0 vs ≥1), comprising covariates consistently ascertained across all three cohorts. Sensitivity analyses evaluating categorical chronic-condition counts (0, 1, 2, or ≥3) and excluding participants with baseline mental illness yielded consistent results (Tables S16 and S17).

The proportional hazards assumption was verified using scaled Schoenfeld residuals, and potential non-linear age effects were evaluated using natural cubic splines with knots at cohort-specific baseline age tertiles (Table S20).

### Subgroup and sensitivity analyses

To evaluate the robustness and clinical specificity of the primary findings, we conducted a prespecified series of sensitivity analyses. First, we refitted the models using complete-case analysis, inverse probability of censoring weighting (IPCW), discrete-time complementary log–log regression across observed survey intervals, and rolling person-period updates across all consecutive wave pairs. Second, to disentangle pharmacological exposure from underlying digestive morbidity in the two cohorts with a harmonised digestive-disease indicator (CHARLS and SHARE; unavailable in HRS), we cross-classified baseline digestive-disease status and medication use, setting untreated digestive disease as the reference group. Third, to account for potential baseline affective vulnerability, we repeated both the baseline and two-wave analyses after excluding participants reporting baseline psychiatric medication use or a history of mental illness (restricting chronic-condition burden to the seven physical conditions) and after additionally adjusting for the continuous baseline depressive-symptom score.

Cohort-specific subgroup models included exposure-by-subgroup terms and Model 4 covariates to estimate stratum-specific HRs and 95% CIs. These estimates were interpreted descriptively (Fig. S4 and Fig. S5).

Three additional analyses evaluated sample selection and exposure stability: (1) computing standardised mean differences in baseline covariates between included participants and those lacking follow-up assessments; (2) re-estimating CHARLS models using the broader gastrointestinal medication definition incorporating traditional Chinese and over-the-counter agents; and (3) performing leave-one-country-out analyses in SHARE by sequentially omitting each participating country (Tables S8–S10 and Figs. S2–S3). All statistical tests were two-sided, with *P* < 0.05 indicating statistical significance.

### Cross-cohort synthesis

Cohort-specific estimates were synthesised using a two-stage random-effects meta-analytic framework to accommodate differences in survey instruments and variable operationalisation across cohorts. For each cohort and exposure contrast, log hazard ratios and corresponding standard errors were pooled using inverse-variance weighted random-effects models with restricted maximum likelihood (REML) estimation of between-cohort variance (τ²), complemented by DerSimonian–Laird estimators in sensitivity analyses. Between-cohort heterogeneity was quantified using Cochran’s *Q* statistic, *I*², and τ², alongside 95% prediction intervals. We additionally computed Hartung–Knapp adjusted confidence intervals, conducted leave-one-cohort-out analyses, and calculated E-values after converting hazard ratios to approximate risk ratios.

### Negative- and positive-control exposures

To evaluate whether the observed associations reflected generic chronic medication use or routine healthcare contact rather than gastrointestinal pharmacotherapy specifically, we repeated the primary baseline analysis (Model 4) across four negative-control medication classes—antihypertensive, lipid-lowering, glucose-lowering, and cardiac medications—and one positive control (psychiatric medication). Negative-control classes were selected as prevalent long-term therapies that share regular prescribing and clinical monitoring in older adults without a gastrointestinal indication. All control analyses used the identical multiply imputed datasets, covariate specification, and cluster-robust Cox proportional hazards framework.

### Competing risk

To account for mortality prior to depressive-symptom onset, we repeated the baseline analysis treating all-cause death during follow-up as a competing risk. Mortality data were retrieved from wave-specific sample files and exit interviews in CHARLS (with unknown exact dates imputed as the midpoint between adjacent survey waves), the RAND HRS Longitudinal File 2020, and the Harmonised SHARE dataset. Subdistribution hazard ratios (sHRs) were estimated using Fine–Gray competing-risk regression models [28], alongside cause-specific Cox models censoring at death. Deaths occurring after the first follow-up assessment without prior threshold crossing were modelled as competing events on the date of death. Potential attrition prior to the first scheduled follow-up was further evaluated using inverse probability of censoring weighted (IPCW) Cox models [29] (Tables S8–S10).

## Results

### Participants and gastrointestinal medication-use patterns

The baseline analysis included 50,467 participants and 16,896 incident depressive-symptom events across CHARLS (6,452 participants; 2,882 events), HRS (12,938 participants; 3,358 events), and SHARE (31,077 participants; 10,656 events) (Fig. 1 and Fig. S1). Median baseline age was 59 years (IQR 55–66) in CHARLS, 65 years (58–74) in HRS, and 64 years (58–71) in SHARE, and women comprised 44.4%, 56.8%, and 52.0% of the respective cohorts. Baseline gastrointestinal medication use was reported by 577 participants (8.9%) in CHARLS, 1,849 (14.3%) in HRS, and 1,680 (5.4%) in SHARE (Table 1A). Over 39,725, 75,700, and 209,762 person-years of follow-up (median 9, 8, and 11 years), incidence rates were 72.6, 44.4, and 50.8 per 1,000 person-years in CHARLS, HRS, and SHARE, respectively.

The two-wave analysis comprised 32,686 participants and 8,069 incident events across CHARLS (4,087 participants; 1,464 events), HRS (9,801 participants; 1,785 events), and SHARE (18,798 participants; 4,820 events). Continued non-use was the predominant pattern (85.7% in CHARLS, 81.8% in HRS, and 91.2% in SHARE), while persistent use at both waves accounted for 4.0%, 9.3%, and 2.6% of participants, respectively (Table 1B).

### Primary associations

After prespecified adjustment (Model 4), baseline gastrointestinal medication use was associated with a higher incidence of depressive symptoms in all three cohorts, with HRs of 1.20 (95% CI 1.07 to 1.36; *P* = 0.002) in CHARLS, 1.35 (1.24 to 1.48; *P* < 0.001) in HRS, and 1.34 (1.24 to 1.44; *P* < 0.001) in SHARE (Table 2A; Fig. 2).

**Figure 2.**
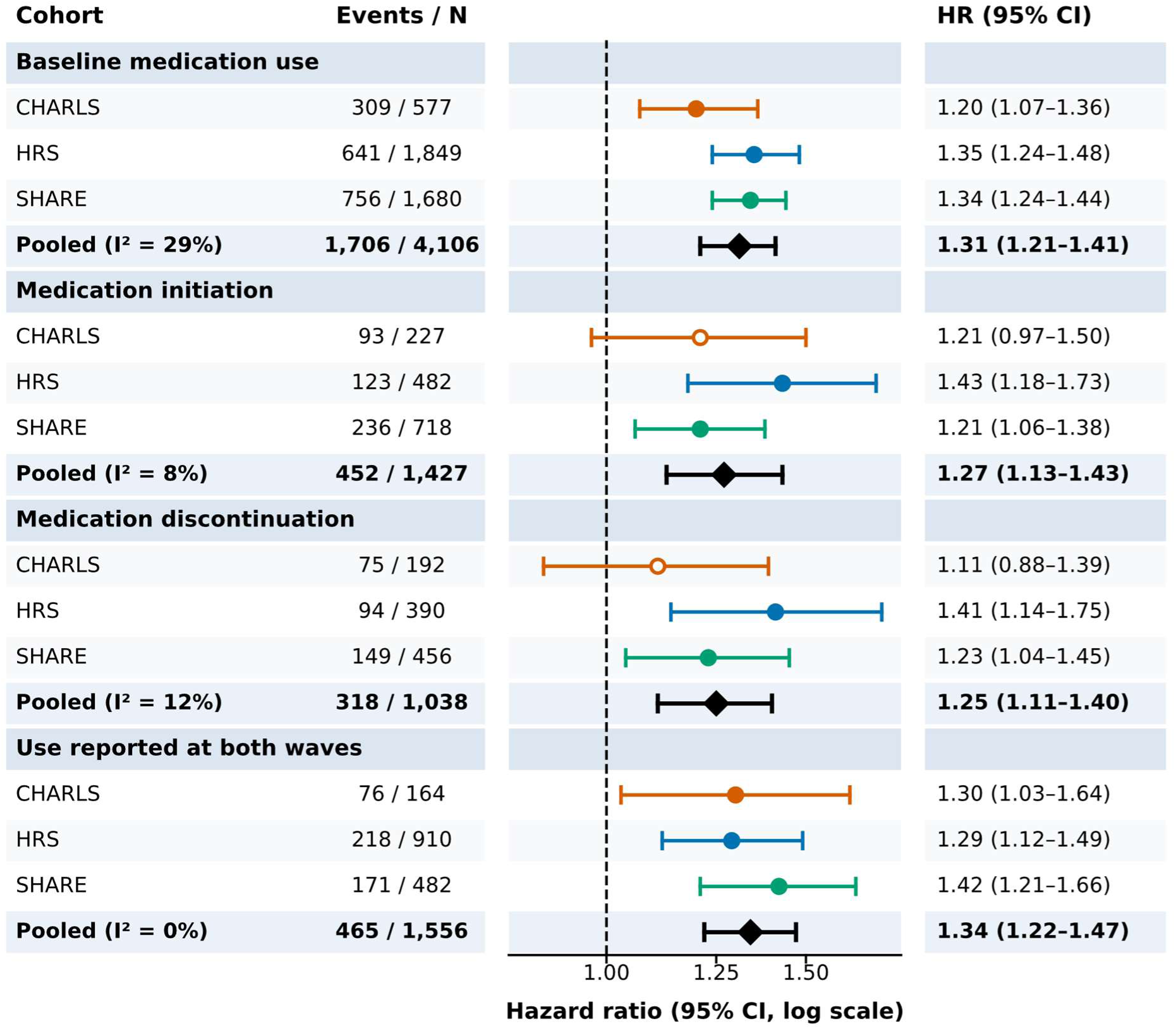
Cohort-specific and pooled associations of gastrointestinal medication use and incident depressive symptoms. Circles show cohort-specific hazard ratios (Model 4) with 95% confidence intervals, coloured by cohort (CHARLS orange, HRS blue, SHARE green); hollow circles indicate confidence intervals that include 1.00; diamonds show random-effects pooled estimates. Rows show baseline medication use versus no use and the three two-wave medication-use patterns against continued non-use, with the number of events and participants within each medication stratum. *I*² is the percentage of variability attributable to between-cohort heterogeneity; the 95% prediction interval for the baseline pooled estimate was 1.16 to 1.47.

**Table 2.**
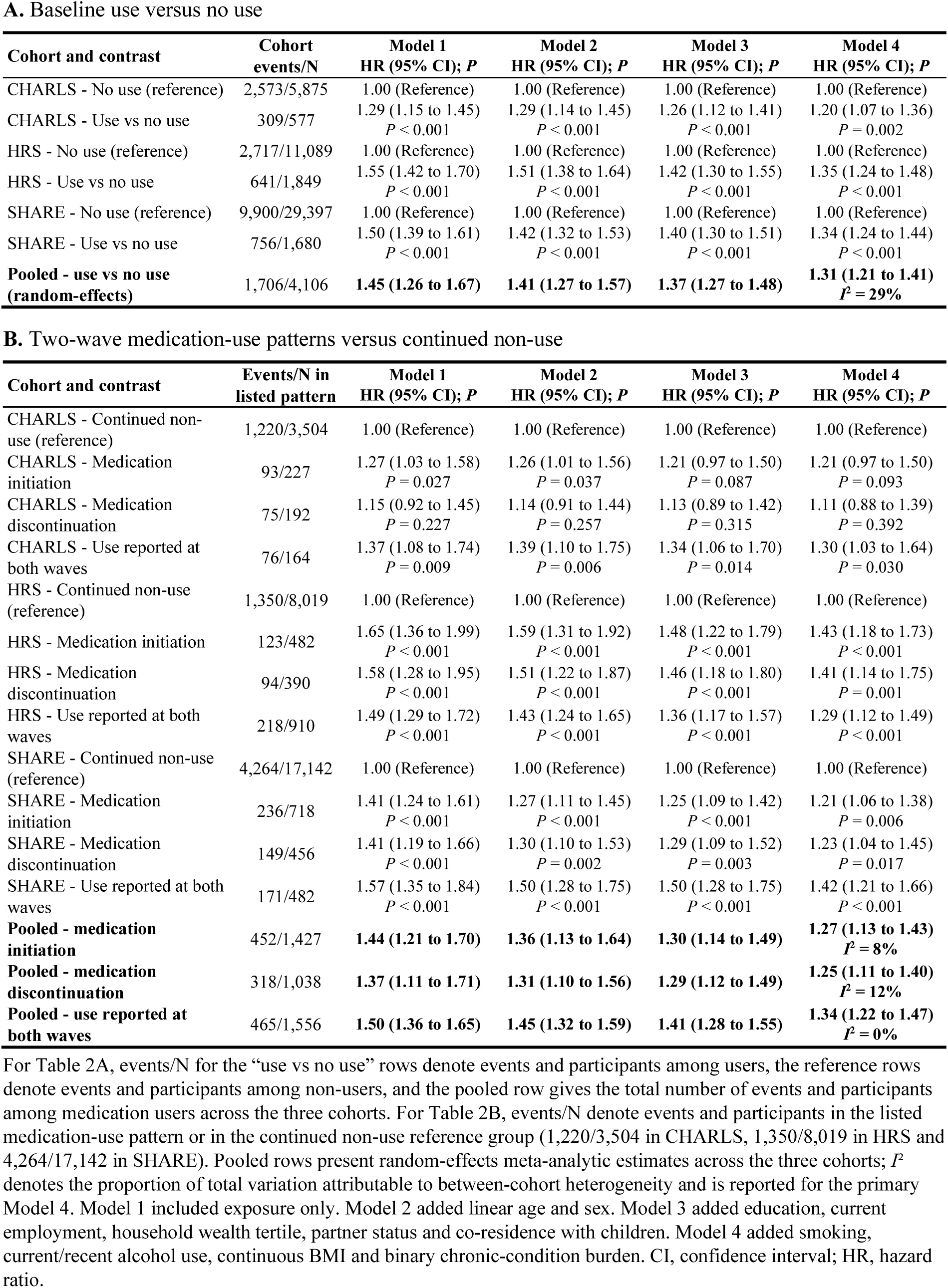
Cohort-specific associations of baseline gastrointestinal medication use and two-wave medication-use patterns with incident depressive symptoms.

**A. Baseline use versus no use**
| Cohort and contrast | Cohort events/N | Model 1 | Model 2 | Model 3 | Model 4 |
| --- | --- | --- | --- | --- | --- |
|  |  | HR (95% CI); *P* | HR (95% CI); *P* | HR (95% CI); *P* | HR (95% CI); *P* |
| CHARLS - No use (reference) | 2,573/5,875 | 1.00 (Reference) | 1.00 (Reference) | 1.00 (Reference) | 1.00 (Reference) |
| CHARLS - Use vs no use | 309/577 | 1.29 (1.15 to 1.45)<br>*P* < 0.001 | 1.29 (1.14 to 1.45)<br>*P* < 0.001 | 1.26 (1.12 to 1.41)<br>*P* < 0.001 | 1.20 (1.07 to 1.36)<br>*P* = 0.002 |
| HRS - No use (reference) | 2,717/11,089 | 1.00 (Reference) | 1.00 (Reference) | 1.00 (Reference) | 1.00 (Reference) |
| HRS - Use vs no use | 641/1,849 | 1.55 (1.42 to 1.70)<br>*P* < 0.001 | 1.51 (1.38 to 1.64)<br>*P* < 0.001 | 1.42 (1.30 to 1.55)<br>*P* < 0.001 | 1.35 (1.24 to 1.48)<br>*P* < 0.001 |
| SHARE - No use (reference) | 9,900/29,397 | 1.00 (Reference) | 1.00 (Reference) | 1.00 (Reference) | 1.00 (Reference) |
| SHARE - Use vs no use | 756/1,680 | 1.50 (1.39 to 1.61)<br>*P* < 0.001 | 1.42 (1.32 to 1.53)<br>*P* < 0.001 | 1.40 (1.30 to 1.51)<br>*P* < 0.001 | 1.34 (1.24 to 1.44)<br>*P* < 0.001 |
| **Pooled - use vs no use (random-effects)** | **1,706/4,106** | **1.45 (1.26 to 1.67)** | **1.41 (1.27 to 1.57)** | **1.37 (1.27 to 1.48)** | **1.31 (1.21 to 1.41)**<br>*I*<sup>2</sup> = 29% |

| Cohort and contrast | Events/N in listed pattern | Model 1 | Model 2 | Model 3 | Model 4 |
| --- | --- | --- | --- | --- | --- |
|  |  | HR (95% CI); *P* | HR (95% CI); *P* | HR (95% CI); *P* | HR (95% CI); *P* |
| CHARLS - Continued non-use (reference) | 1,220/3,504 | 1.00 (Reference) | 1.00 (Reference) | 1.00 (Reference) | 1.00 (Reference) |
| CHARLS - Medication initiation | 93/227 | 1.27 (1.03 to 1.58)<br>*P* = 0.027 | 1.26 (1.01 to 1.56)<br>*P* = 0.037 | 1.21 (0.97 to 1.50)<br>*P* = 0.087 | 1.21 (0.97 to 1.50)<br>*P* = 0.093 |
| CHARLS - Medication discontinuation | 75/192 | 1.15 (0.92 to 1.45)<br>*P* = 0.227 | 1.14 (0.91 to 1.44)<br>*P* = 0.257 | 1.13 (0.89 to 1.42)<br>*P* = 0.315 | 1.11 (0.88 to 1.39)<br>*P* = 0.392 |
| CHARLS - Use reported at both waves | 76/164 | 1.37 (1.08 to 1.74)<br>*P* = 0.009 | 1.39 (1.10 to 1.75)<br>*P* = 0.006 | 1.34 (1.06 to 1.70)<br>*P* = 0.014 | 1.30 (1.03 to 1.64)<br>*P* = 0.030 |
| HRS - Continued non-use (reference) | 1,350/8,019 | 1.00 (Reference) | 1.00 (Reference) | 1.00 (Reference) | 1.00 (Reference) |
| HRS - Medication initiation | 123/482 | 1.65 (1.36 to 1.99)<br>*P* < 0.001 | 1.59 (1.31 to 1.92)<br>*P* < 0.001 | 1.48 (1.22 to 1.79)<br>*P* < 0.001 | 1.43 (1.18 to 1.73)<br>*P* < 0.001 |
| HRS - Medication discontinuation | 94/390 | 1.58 (1.28 to 1.95)<br>*P* < 0.001 | 1.51 (1.22 to 1.87)<br>*P* < 0.001 | 1.46 (1.18 to 1.80)<br>*P* < 0.001 | 1.41 (1.14 to 1.75)<br>*P* = 0.001 |
| HRS - Use reported at both waves | 218/910 | 1.49 (1.29 to 1.72)<br>*P* < 0.001 | 1.43 (1.24 to 1.65)<br>*P* < 0.001 | 1.36 (1.17 to 1.57)<br>*P* < 0.001 | 1.29 (1.12 to 1.49)<br>*P* < 0.001 |
| SHARE - Continued non-use (reference) | 4,264/17,142 | 1.00 (Reference) | 1.00 (Reference) | 1.00 (Reference) | 1.00 (Reference) |
| SHARE - Medication initiation | 236/718 | 1.41 (1.24 to 1.61)<br>*P* < 0.001 | 1.27 (1.11 to 1.45)<br>*P* < 0.001 | 1.25 (1.09 to 1.42)<br>*P* < 0.001 | 1.21 (1.06 to 1.38)<br>*P* = 0.006 |
| SHARE - Medication discontinuation | 149/456 | 1.41 (1.19 to 1.66)<br>*P* < 0.001 | 1.30 (1.10 to 1.53)<br>*P* = 0.002 | 1.29 (1.09 to 1.52)<br>*P* = 0.003 | 1.23 (1.04 to 1.45)<br>*P* = 0.017 |
| SHARE - Use reported at both waves | 171/482 | 1.57 (1.35 to 1.84)<br>*P* < 0.001 | 1.50 (1.28 to 1.75)<br>*P* < 0.001 | 1.50 (1.28 to 1.75)<br>*P* < 0.001 | 1.42 (1.21 to 1.66)<br>*P* < 0.001 |
| **Pooled - medication initiation** | **452/1,427** | **1.44 (1.21 to 1.70)** | **1.36 (1.13 to 1.64)** | **1.30 (1.14 to 1.49)** | **1.27 (1.13 to 1.43)**<br>*I*<sup>2</sup> = 8% |
| **Pooled - medication discontinuation** | **318/1,038** | **1.37 (1.11 to 1.71)** | **1.31 (1.10 to 1.56)** | **1.29 (1.12 to 1.49)** | **1.25 (1.11 to 1.40)**<br>*I*<sup>2</sup> = 12% |
| **Pooled - use reported at both waves** | **465/1,556** | **1.50 (1.36 to 1.65)** | **1.45 (1.32 to 1.59)** | **1.41 (1.28 to 1.55)** | **1.34 (1.22 to 1.47)**<br>*I*<sup>2</sup> = 0% |

Compared with continued non-use, medication initiation was associated with HRs of 1.21 (95% CI 0.97 to 1.50; *P* = 0.093) in CHARLS, 1.43 (1.18 to 1.73; *P* < 0.001) in HRS, and 1.21 (1.06 to 1.38; *P* = 0.006) in SHARE. For medication discontinuation, the corresponding HRs were 1.11 (0.88 to 1.39; *P* = 0.392), 1.41 (1.14 to 1.75; *P* = 0.001), and 1.23 (1.04 to 1.45; *P* = 0.017).

Persistent medication use reported at both waves showed the strongest and most consistent association across cohorts: HR 1.30 (95% CI 1.03 to 1.64; *P* = 0.030) in CHARLS, 1.29 (1.12 to 1.49; *P* < 0.001) in HRS, and 1.42 (1.21 to 1.66; *P* < 0.001) in SHARE (Table 2B; Fig. 2).

In random-effects meta-analyses, pooled summary hazard ratios were 1.31 (95% CI 1.21 to 1.41; *I*² = 29%) for baseline use and 1.34 (1.22 to 1.47; *I*² = 0%) for persistent use, with pooled estimates of 1.27 (1.13 to 1.43; *I*² = 8%) for initiation and 1.25 (1.11 to 1.40; *I*² = 12%) for discontinuation (Table S12).

### Sensitivity and secondary analyses

Progressive adjustment for lifestyle factors, BMI, and chronic-condition burden in Model 4 modestly attenuated effect sizes relative to Model 3, but baseline gastrointestinal medication use and persistent two-wave use remained significantly associated with incident depressive symptoms across all three cohorts. Medication initiation and discontinuation also remained significantly associated with incident depressive symptoms in HRS and SHARE, whereas estimates in the smaller CHARLS transition groups were imprecise.

Complete-case, IPCW, and discrete-time complementary log–log models yielded estimates consistent with the primary multiple-imputation analyses across all cohorts (Tables S8–S10 and Fig. S2). Scaled Schoenfeld residuals confirmed that the proportional hazards assumption was satisfied for the exposure terms in every cohort (*P* = 0.28 in CHARLS, *P* = 0.06 in HRS, and *P* = 0.66 in SHARE); stratifying on covariates showing minor non-proportionality in global tests (*P* = 0.021, *P* < 0.001, and *P* < 0.001, respectively) left exposure estimates unchanged.

Excluding participants reporting baseline psychiatric medication use yielded consistent estimates in CHARLS (HR 1.21, 95% CI 1.07 to 1.36), HRS (1.26, 1.13 to 1.41), and SHARE (1.32, 1.22 to 1.43; pooled HR 1.28, 95% CI 1.21 to 1.35; *I*² = 0%), and further excluding those with a diagnosed mental illness gave 1.21 (1.07 to 1.36), 1.26 (1.12 to 1.41), and 1.33 (1.22 to 1.44), respectively (Table S16). Baseline subthreshold depressive-symptom scores were modestly higher among medication users than non-users across cohorts (mean 5.10 vs 4.35 in CHARLS, 0.73 vs 0.53 in HRS, and 1.68 vs 1.33 in SHARE; Tables S2–S7). After further adjusting for the continuous baseline depressive-symptom score, the pooled association remained robust (pooled HR 1.21, 95% CI 1.14 to 1.27; *I*² = 0%), with cohort-specific HRs of 1.12 (95% CI 1.00 to 1.27) in CHARLS, 1.23 (1.12 to 1.34) in HRS, and 1.23 (1.14 to 1.33) in SHARE. Categorising chronic-condition burden as 0, 1, 2, or ≥3 conditions likewise yielded consistent estimates (1.19 [1.06 to 1.34], 1.24 [1.14 to 1.36], and 1.26 [1.17 to 1.36], respectively; Table S17).

In the two cohorts with a harmonised self-reported digestive-disease diagnosis (CHARLS and SHARE), using participants with untreated digestive disease as the reference group (HR 1.00), those with neither digestive disease nor medication had a significantly lower risk of incident depressive symptoms (HR 0.87, 95% CI 0.77 to 0.98 in CHARLS; HR 0.82, 95% CI 0.75 to 0.89 in SHARE; corresponding to reciprocal HRs of 1.15 [95% CI 1.02 to 1.30] and 1.22 [95% CI 1.12 to 1.33] for untreated digestive disease versus no disease). Notably, among participants with digestive disease, concurrent gastrointestinal medication use was associated with a further elevation in risk compared with untreated digestive disease (HR 1.21, 95% CI 1.03 to 1.42 in SHARE; HR 1.14, 95% CI 0.96 to 1.35 in CHARLS; pooled HR 1.18, 95% CI 1.05 to 1.32; *I*² = 0%; Table 3 and Table S18). The two-wave persistent-use associations were similarly robust after additional adjustment for the continuous baseline symptom score (HR 1.20 [95% CI 0.95 to 1.52] in CHARLS, 1.20 [1.04 to 1.39] in HRS, and 1.30 [1.11 to 1.53] in SHARE; pooled HR 1.24, 95% CI 1.12 to 1.37; *I*² = 0%) and after excluding baseline psychiatric medication users (HR 1.31 [1.03 to 1.65] in CHARLS, 1.18 [0.99 to 1.41] in HRS, and 1.39 [1.18 to 1.64] in SHARE; pooled HR 1.30, 95% CI 1.16 to 1.46; *I*² = 13%; Table 3 and Table S19). Modelling age with natural cubic splines yielded virtually identical estimates across all cohorts (Table S20).

**Table 3.**
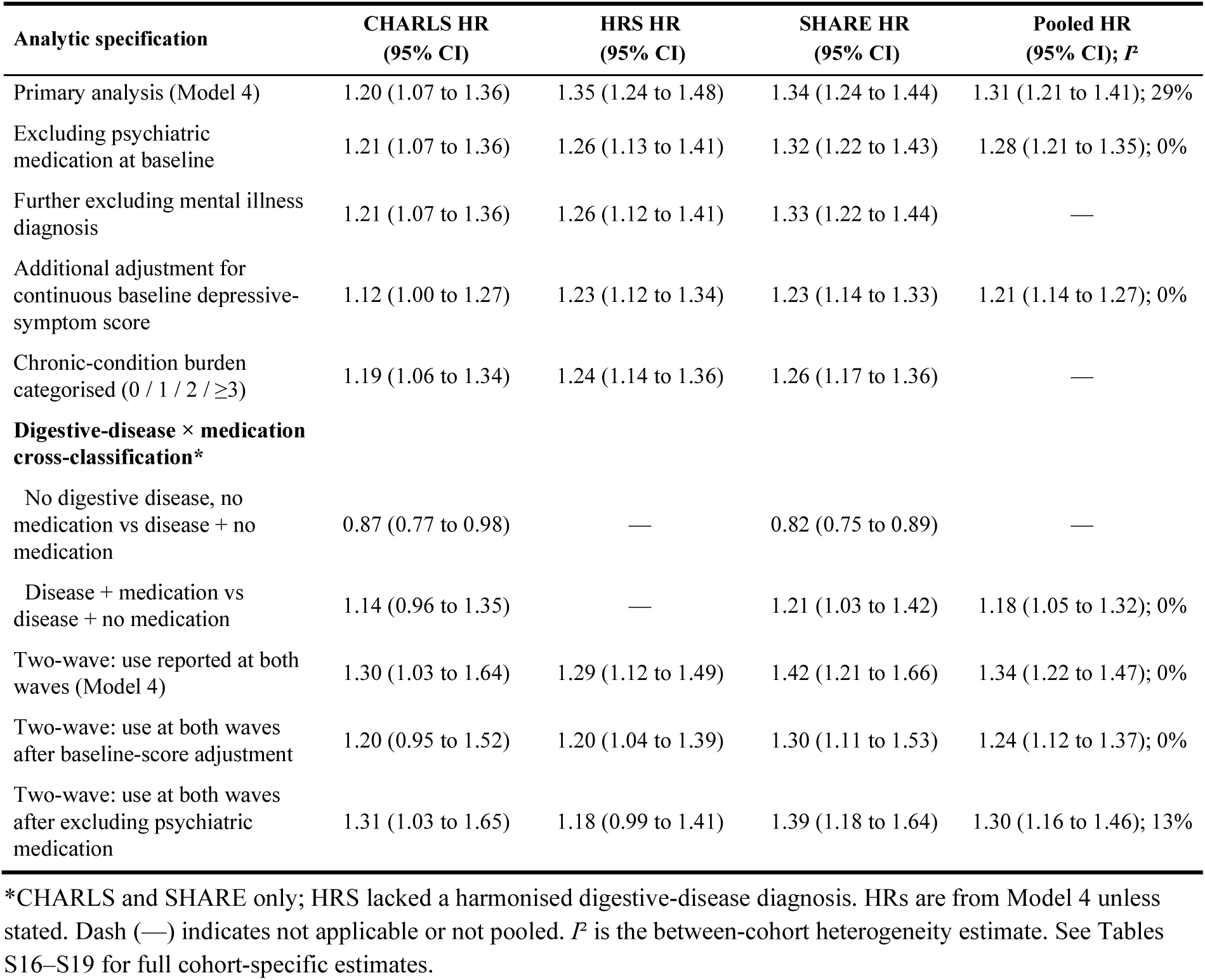
Sensitivity and specificity analyses of gastrointestinal medication use and incident depressive symptoms, by cohort and pooled estimate.

Baseline characteristics of included participants and those without follow-up assessments are compared in Fig. S6; included participants in CHARLS and HRS were modestly younger and more frequently employed, whereas differences across all covariates in SHARE were minimal.

Applying the broader CHARLS exposure definition incorporating traditional Chinese and over-the-counter medications yielded a baseline HR of 1.14 (95% CI 1.02 to 1.28) and an HR of 1.24 (1.00 to 1.54) for persistent use at both waves. In SHARE, leave-one-country-out analyses confirmed that no single country drove the associations, with HRs ranging from 1.30 to 1.36 for baseline use, 1.17 to 1.24 for initiation, 1.18 to 1.27 for discontinuation, and 1.36 to 1.46 for persistent use (Fig. S3).

Time-updated rolling person-period analyses (encompassing 7,577 intervals in CHARLS, 22,390 in HRS, and 27,129 in SHARE) reinforced the primary two-wave findings across all cohorts, including a statistically significant association for medication discontinuation in CHARLS (HR 1.32, 95% CI 1.02 to 1.71; *P* = 0.032) when incorporating all repeated exposure intervals (Table S11).

Stratum-specific estimates were generally consistent in direction across demographic, socioeconomic, lifestyle, and health-burden subgroups (Table S13 and Figs. S4–S5). Across 30 formal exposure-by-subgroup interaction tests in the baseline analysis, only educational attainment in HRS reached nominal statistical significance (*P* for interaction = 0.015), where the association was stronger among participants with low education (HR 1.66, 95% CI 1.38 to 2.00) than among those with middle or high education (HR 1.28, 95% CI 1.16 to 1.41), accompanied by a similar numerical gradient across household wealth tertiles (HR 1.46, 1.32, and 1.21 from lowest to highest tertile). Given the number of subgroup comparisons conducted, these stratum-specific differences were interpreted descriptively. Full stratum-specific estimates with 95% CIs are provided in Table S13.

### Consistency across cohorts

Meta-analytic synthesis confirmed a consistent association between baseline gastrointestinal medication use and incident depressive symptoms across the three cohorts (pooled HR 1.31, 95% CI 1.21 to 1.41; *I*² = 29%, *P* for heterogeneity = 0.24; 95% prediction interval 1.16 to 1.47). Among two-wave patterns, persistent use at both waves demonstrated the strongest and most homogeneous association (pooled HR 1.34, 95% CI 1.22 to 1.47; *I*² = 0%), followed by initiation (1.27, 1.13 to 1.43; *I*² = 8%) and discontinuation (1.25, 1.11 to 1.40; *I*² = 12%) (Fig. 2 and Table S12). Leave-one-cohort-out analyses yielded stable summary estimates for baseline use (range 1.28 to 1.34) and persistent use (range 1.29 to 1.38), and Hartung–Knapp confidence intervals excluded unity for both contrasts (1.31 [1.11 to 1.54] and 1.34 [1.08 to 1.65], respectively). Progressive covariate adjustment reduced between-cohort heterogeneity for baseline use from *I*² = 69% in Model 1 to 29% in Model 4 while maintaining *I*² = 0% for persistent use across all models. E-values for the primary pooled estimates ranged from 1.60 to 1.75 (lower 95% CI limits 1.36 to 1.55).

### Negative- and positive-control exposures

Antihypertensive medication (HR 0.90, 95% CI 0.81 to 1.00 in CHARLS; 1.00, 0.93 to 1.08 in HRS; 0.96, 0.91 to 1.00 in SHARE; pooled HR 0.96, 95% CI 0.91 to 1.01) and lipid-lowering medication (1.08, 0.90 to 1.31; 1.00, 0.93 to 1.07; 1.04, 1.00 to 1.09; pooled HR 1.03, 95% CI 0.99 to 1.07) were not associated with incident depressive symptoms in any cohort. Cardiac medication was associated with higher risk in HRS (1.24, 1.12 to 1.36) and SHARE (1.27, 1.20 to 1.34), and glucose-lowering medication in SHARE (1.16, 1.09 to 1.24). Psychiatric medication (positive control) showed the expected strong association in HRS (2.10, 1.93 to 2.29) and SHARE (1.89, 1.72 to 2.06; pooled HR 1.99, 95% CI 1.79 to 2.22), whereas only 15 participants reported baseline psychiatric medication use in CHARLS (Fig. 3 and Table S14).

**Figure 3.**
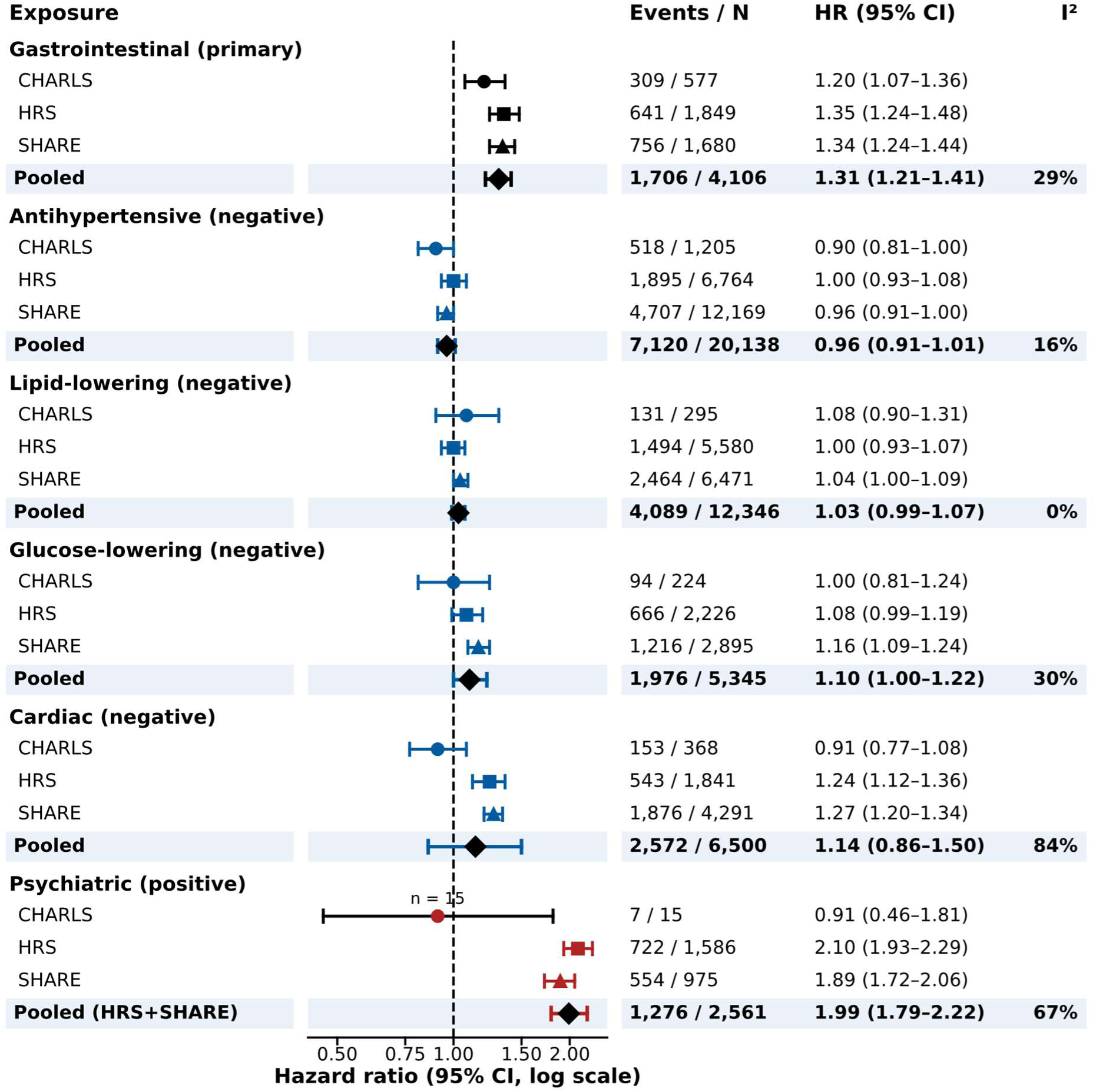
Specificity of the association: negative- and positive-control exposures. Hazard ratios (Model 4, 95% confidence intervals) for gastrointestinal medication (primary exposure), four negative-control medication classes and psychiatric medication (positive control) in each cohort and pooled across cohorts; negative-control estimates are shown in blue-grey, the primary exposure in black and the positive control in red. Diamonds show random-effects pooled estimates and *I*² is shown for each pooled estimate. Events/N are observed incident events and participants in each exposure group (before imputation), not the total cohort events. Because only 15 participants in CHARLS reported baseline psychiatric medication use (labelled *n* = 15), the pooled positive-control estimate combines HRS and SHARE (HR 1.99, 95% CI 1.79–2.22; *I*² = 67%).

### Competing risk

During follow-up, 172 (2.7%) participants in CHARLS, 1,353 (10.5%) in HRS and 2,232 (7.2%) in SHARE died before the end of the follow-up window. Treating death as a competing event did not materially change the estimates: subdistribution hazard ratios were 1.22 (95% CI 1.08 to 1.37), 1.35 (1.24 to 1.48) and 1.33 (1.24 to 1.44), and cause-specific hazard ratios 1.20 (1.07 to 1.36), 1.36 (1.24 to 1.48) and 1.34 (1.24 to 1.44) in CHARLS, HRS and SHARE, respectively (Table S15).

## Discussion

In this multicohort study across China, the United States, and 16 European countries, baseline gastrointestinal medication use was consistently associated with increased risk of incident depressive symptoms (pooled HR 1.31, 95% CI 1.21–1.41; *I*² = 29%). Furthermore, persistent medication use across consecutive survey waves demonstrated the most robust and homogeneous association (pooled HR 1.34, 95% CI 1.22–1.47; *I*² = 0%). These findings demonstrate that gastrointestinal pharmacotherapy in older adults is reliably linked to subsequent depressive symptoms across diverse healthcare systems, highlighting medication review as a key clinical touchpoint for mental health assessment.

Prior literature examining gastrointestinal medications and mood has yielded disparate observations, largely constrained by cross-sectional designs and single-population settings. Observational analyses in US national cohorts identified higher depression prevalence among users of prescription medications with potential depressogenic effects, including proton pump inhibitors [11–13]. Bidirectional relationships between gastro-oesophageal reflux and affective disorders are well documented [16, 17], and gastrointestinal symptoms frequently co-occur with psychological distress [3–6]. Several biological mechanisms provide plausible explanations for our longitudinal findings. Gastric acid suppression, particularly with long-term proton pump inhibitor use, can induce profound alterations in the gut microbiome, shift short-chain fatty acid profiles, and disrupt the bidirectional gut–brain axis [9, 10, 20].

Moreover, hypochlorhydria impairs the absorption of essential micronutrients, including vitamin B12 and magnesium, which play established roles in neurotransmitter synthesis and mood regulation. Concurrently, systemic low-grade inflammation associated with chronic digestive pathology may facilitate neuroinflammatory cascades that heighten vulnerability to depressive episodes.

Clinical trials and short-term prospective studies have reported symptomatic relief or modest cognitive-affective improvements following initial treatment of severe reflux disease [18].

However, in broader real-world populations, inappropriate and long-term use of proton pump inhibitors without documented clinical indications is widespread [8]. In this context, self-reported gastrointestinal medication use may capture both pharmacological effects and the burden of chronic digestive disease [9, 10, 19]. Rather than viewing drug effects and disease morbidity as mutually exclusive, our longitudinal data indicate that sustained pharmacotherapy identifies individuals experiencing both ongoing digestive distress and prolonged drug exposure.

Two key analytical findings underscore the specificity and independence of our observations. First, although baseline depressive symptom scores were slightly higher among medication users, the association remained robust and homogeneous after adjusting for continuous baseline subthreshold depressive symptoms (pooled HR 1.21, 95% CI 1.14–1.27; *I*² = 0%; Table S17), demonstrating that the risk elevation is not an artefact of proximity to the diagnostic threshold. Second, in digestive disease–stratified analyses in CHARLS and SHARE, gastrointestinal medication use was associated with an incremental risk increase beyond untreated digestive disease (pooled HR 1.18, 95% CI 1.05–1.32; *I*² = 0%; Table S18). Together with the stability of estimates across levels of multimorbidity, these findings indicate that while underlying digestive morbidity contributes to psychological vulnerability, gastrointestinal pharmacotherapy confers an additional, independent risk.

The attenuated estimates for two-wave medication initiation and discontinuation in CHARLS reflect lower statistical power in the smaller transition subgroups and a more specific exposure definition tied to physician-diagnosed conditions. Across all three cohorts, persistent medication use across consecutive waves yielded the most consistent and robust risk elevation, highlighting sustained exposure or chronic underlying disease as the primary driver of subsequent depressive risk.

The specificity of our findings is reinforced by negative- and positive-control exposures. Antihypertensive and lipid-lowering therapies—widely used chronic medications with frequent clinical monitoring—showed null associations with incident depression across all cohorts (pooled HRs 0.96 and 1.03, respectively). This confirms that our primary findings are not attributable to general prescription reporting, healthcare contact frequency, or health-seeking behaviour. In contrast, psychiatric medications (positive control) demonstrated strong, expected associations (pooled HR 1.99). Additionally, subgroup analyses in HRS indicated more pronounced associations among socioeconomically disadvantaged adults. This pattern aligns with World Health Organization guidelines on Integrated Care for Older People (ICOPE) [30], which emphasise comprehensive, person-centred assessment for vulnerable older adults experiencing multimorbidity and polypharmacy.

Our study has multiple methodological strengths. We utilised harmonised prospective data from three large national ageing cohorts spanning diverse healthcare systems and socioeconomic contexts. The longitudinal design ensured that medication assessment preceded depressive symptom ascertainment among individuals initially free of clinically significant symptoms. The evaluation of two-wave medication patterns distinguished persistent users from those initiating or discontinuing therapy. Furthermore, we employed rigorous analytical strategies, including multiple imputation, negative-control exposures, competing-risk modelling for mortality, and leave-one-cohort-out validation, ensuring high internal and external validity.

Several limitations should be considered. First, medication use was ascertained via self-report, and specific dosages, active chemical entities (e.g., proton pump inhibitors vs H_2_ receptor antagonists), and exact treatment durations were unavailable in the survey files; such non-differential exposure misclassification would typically bias estimates toward the null. Second, medication status between survey waves was unobserved, so two-wave patterns reflect status at biennial or triennial interviews rather than continuous daily adherence. Third, although Fine–Gray competing-risk and inverse-probability-weighted models confirmed that mortality and follow-up attrition did not materially affect the estimates, incident depressive symptoms could only be assessed among survivors attending follow-up interviews. Fourth, while models adjusted for harmonized sociodemographic, behavioural, and chronic health covariates, residual confounding by unmeasured clinical severity or lifestyle factors (such as physical activity or functional status) cannot be entirely excluded. Finally, cohort-specific symptom scales (CES-D-10, CES-D-8, and EURO-D) preclude direct pooling of raw scores, although two-stage random-effects synthesis demonstrated high cross-national consistency in relative risk estimates.

These findings have direct clinical and public health relevance. Older adults presenting with gastrointestinal complaints or receiving gastrointestinal pharmacotherapy represent an accessible target population for routine mental health assessment. Rather than suggesting that gastrointestinal medications should be withheld when clinically indicated, our results suggest that medication reviews provide a valuable clinical window to identify older adults at elevated risk of depressive symptoms. Integrating brief, routine mood screening into primary care management of older adults on long-term gastrointestinal therapy could facilitate timely psychosocial support and promote healthy cognitive and emotional longevity.

## Conclusions

Across three national ageing cohorts, baseline and persistent two-wave gastrointestinal medication use were consistently associated with an elevated incidence of depressive symptoms, independent of baseline subthreshold symptoms, underlying digestive morbidity, and competing mortality. Routine clinical review of long-term gastrointestinal pharmacotherapy provides a practical opportunity for proportionate affective screening in older adults, while prescribing decisions should remain guided by established gastrointestinal indications.

## Data Availability

All data analysed in the present study are available online to registered researchers subject to the data-access requirements of the respective cohort repositories: the China Health and Retirement Longitudinal Study (CHARLS, https://charls.pku.edu.cn/), the Health and Retirement Study (HRS, https://hrsdata.isr.umich.edu/), the Survey of Health, Ageing and Retirement in Europe (SHARE, https://share-eric.eu/data/data-access), and the Gateway to Global Aging Data (https://g2aging.org/).

https://hrsdata.isr.umich.edu/

https://share-eric.eu/data/data-access

## Abbreviations

BMI: body mass index
CES-D: Center for Epidemiologic Studies Depression scale
CHARLS: China Health and Retirement Longitudinal Study
CI: confidence interval
EURO-D: European Union initiative to compare symptoms of depression scale
HR: hazard ratio
HRS: Health and Retirement Study
IPCW: inverse probability of censoring weighting
IQR: interquartile range
MICE: multiple imputation by chained equations
REML: restricted maximum likelihood
SD: standard deviation
SHARE: Survey of Health, Ageing and Retirement in Europe
sHR: subdistribution hazard ratio
STROBE: Strengthening the Reporting of Observational Studies in Epidemiology

## Acknowledgements

We thank the participants, interviewers, investigators and data-management teams of the CHARLS, the HRS, and the SHARE for making these cohort resources available for scientific research. During the preparation of this work, the authors used ChatGPT (OpenAI) to improve language and readability. After using this tool, the authors reviewed and edited the content as needed and take full responsibility for the content of the publication. The CHARLS, HRS and SHARE harmonised data files were made available through the Gateway to Global Aging Data.

## Authors’ contributions

DL and SW curated the data, conducted the statistical analyses and prepared the initial draft of the manuscript. TT contributed to the interpretation of the findings and critically revised the manuscript. SW contributed to the critical revision of the manuscript and overall supervision. All authors reviewed and approved the final manuscript. SW is the guarantor and accepts full responsibility for the finished work and the conduct of the study, had access to the data and controlled the decision to publish.

## Funding

This study was supported by the Shenzhen Science and Technology Program (JCYJ20240813114815021 and JCYJ20250604185328037). The funder had no role in study design, data collection, analysis, interpretation, writing or the decision to submit. The HRS is funded by the US National Institute on Aging (NIA grant U01AG009740).

## Availability of data and materials

CHARLS, HRS and SHARE data are available to registered researchers subject to the access and terms-of-use requirements of the respective cohort repositories: https://charls.pku.edu.cn/; https://hrsdata.isr.umich.edu/; and https://share-eric.eu/data/data-access.

## Declarations

### Ethics approval and consent to participate

The present study was a secondary analysis of de-identified data and involved no direct contact with participants; no additional ethics approval or registration was required at the authors’ institution. The CHARLS protocol was approved by the Institutional Review Board at Peking University (IRB00001052-11015). The HRS protocol was approved by the University of Michigan Health Sciences/Behavioral Sciences Institutional Review Board. The SHARE protocol was approved by the Ethics Committee of the University of Mannheim/the Max Planck Society. Participants provided informed consent in accordance with the approved study procedures.

### Consent for publication

Not applicable.

### Competing interests

The authors declare that they have no competing interests.

## Supplementary Information

Supplementary tables (Tables S1–S22), supplementary figures (Figures S1–S6), and the STROBE checklist are provided in Additional file 1 and Additional file 2.

### Additional file 1: Supplementary tables and figures

**Table S1.**
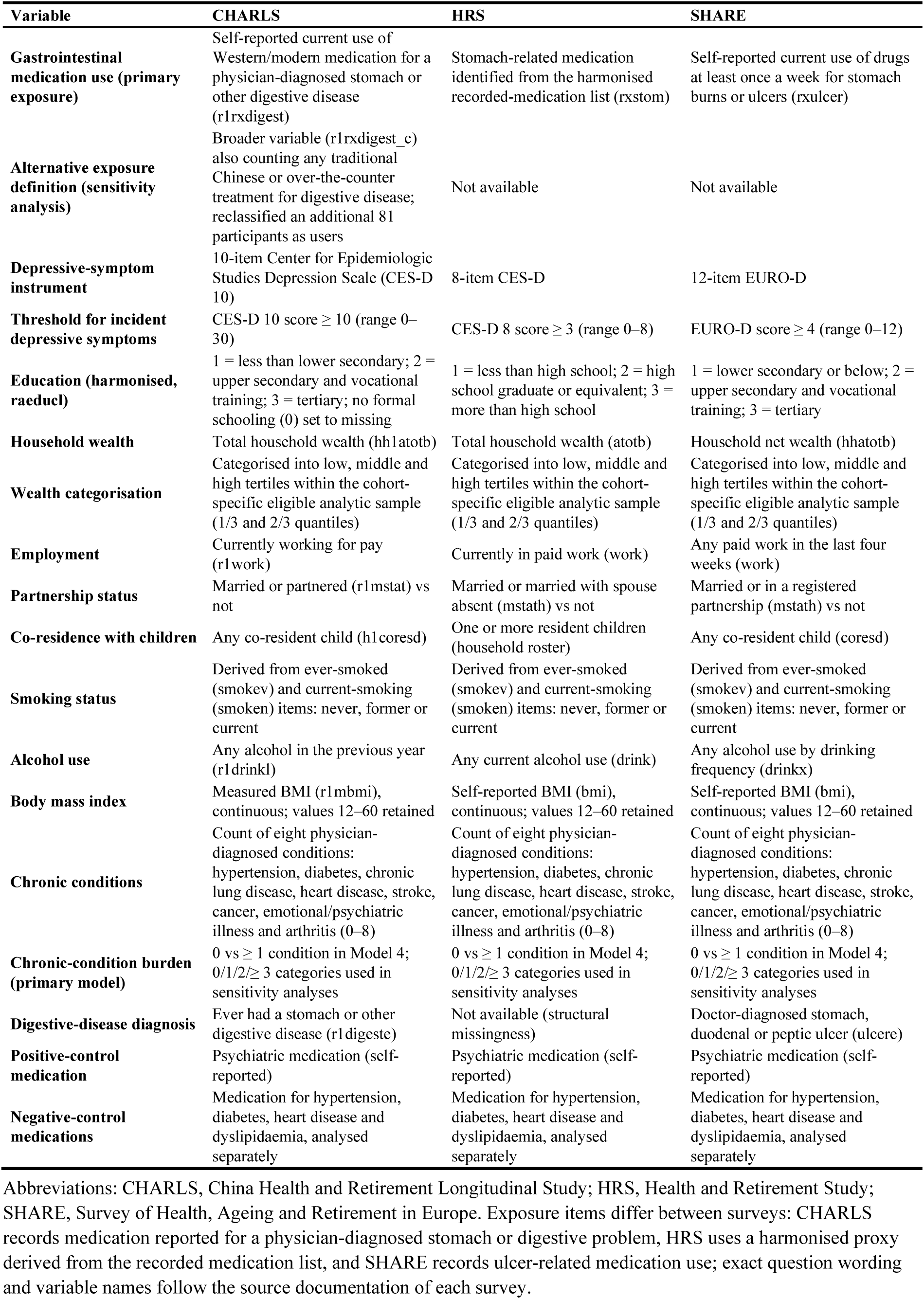
Harmonised definitions of key variables in the three cohorts.

**Table S2.** Baseline characteristics by gastrointestinal medication use in CHARLS.

| Characteristic | No use (*n*=5,875) | Use (*n*=577) |
| --- | --- | --- |
| Age, years, median (IQR) | 59 (55–66) | 59 (55–64) |
| **Sex, *n* (%)** |  |  |
| Male | 3,283 (55.9) | 307 (53.2) |
| Female | 2,592 (44.1) | 270 (46.8) |
| **Education, *n* (%)** |  |  |
| Low | 5,039 (85.8) | 513 (88.9) |
| Middle | 698 (11.9) | 57 (9.9) |
| High | 138 (2.3) | 7 (1.2) |
| **Currently employed, *n* (%)** |  |  |
| No | 2,256 (38.9) | 175 (30.8) |
| Yes | 3,541 (61.1) | 394 (69.2) |
| **Household wealth tertile, *n* (%)** |  |  |
| Low | 1,377 (32.8) | 135 (32.7) |
| Middle | 1,412 (33.7) | 162 (39.2) |
| High | 1,406 (33.5) | 116 (28.1) |
| **Married/partnered, *n* (%)** |  |  |
| No | 634 (10.8) | 47 (8.1) |
| Yes | 5,241 (89.2) | 530 (91.9) |
| **Co-residence with children, *n* (%)** |  |  |
| No | 2,546 (44.3) | 233 (41.0) |
| Yes | 3,197 (55.7) | 335 (59.0) |
| **Smoking status, *n* (%)** |  |  |
| Never | 3,286 (56.0) | 329 (57.0) |
| Former | 595 (10.1) | 54 (9.4) |
| Current | 1,991 (33.9) | 194 (33.6) |
| **Current drinking, *n* (%)** |  |  |
| No | 3,710 (63.1) | 403 (69.8) |
| Yes | 2,165 (36.9) | 174 (30.2) |
| Body mass index, kg/m<sup>2</sup>, mean (SD) | 23.60 (3.77) | 23.08 (3.81) |
| **Chronic-condition burden, *n* (%)** |  |  |
| 0 | 2,640 (45.7) | 205 (36.6) |
| ≥1 | 3,143 (54.3) | 355 (63.4) |
| Depressive-symptom score at risk origin, mean (SD) | 4.35 (2.77) | 5.10 (2.79) |

Abbreviations: CHARLS, China Health and Retirement Longitudinal Study; IQR, interquartile range; SD, standard deviation.

**Table S3.** Baseline characteristics by gastrointestinal medication use in HRS.

| Characteristic | No use (*n*=11,089) | Use (*n*=1,849) |
| --- | --- | --- |
| Age, years, median (IQR) | 65 (58–74) | 69 (60–76) |
| **Sex, *n* (%)** |  |  |
| Male | 4,887 (44.1) | 699 (37.8) |
| Female | 6,202 (55.9) | 1,150 (62.2) |
| **Education, *n* (%)** |  |  |
| Low | 1,572 (14.2) | 320 (17.3) |
| Middle | 6,578 (59.3) | 1,122 (60.7) |
| High | 2,936 (26.5) | 407 (22.0) |
| **Currently employed, *n* (%)** |  |  |
| No | 5,868 (53.0) | 1,267 (68.5) |
| Yes | 5,211 (47.0) | 582 (31.5) |
| **Household wealth tertile, *n* (%)** |  |  |
| Low | 3,598 (32.4) | 688 (37.2) |
| Middle | 3,682 (33.2) | 634 (34.3) |
| High | 3,809 (34.3) | 527 (28.5) |
| **Married/partnered, *n* (%)** |  |  |
| No | 4,079 (36.8) | 722 (39.0) |
| Yes | 7,010 (63.2) | 1,127 (61.0) |
| **Co-residence with children, *n* (%)** |  |  |
| No | 7,976 (71.9) | 1,419 (76.7) |
| Yes | 3,112 (28.1) | 430 (23.3) |
| **Smoking status, *n* (%)** |  |  |
| Never | 5,124 (46.4) | 764 (41.6) |
| Former | 4,522 (41.0) | 879 (47.8) |
| Current | 1,389 (12.6) | 194 (10.6) |
| **Current drinking, *n* (%)** |  |  |
| No | 4,482 (40.4) | 896 (48.5) |
| Yes | 6,604 (59.6) | 953 (51.5) |
| Body mass index, kg/m<sup>2</sup>, mean (SD) | 28.33 (5.74) | 29.32 (5.93) |
| **Chronic-condition burden, *n* (%)** |  |  |
| 0 | 2,145 (19.3) | 87 (4.7) |
| ≥1 | 8,944 (80.7) | 1,762 (95.3) |
| Depressive-symptom score at risk origin, mean (SD) | 0.53 (0.71) | 0.73 (0.78) |

Abbreviations: HRS, Health and Retirement Study; IQR, interquartile range; SD, standard deviation.

**Table S4.**
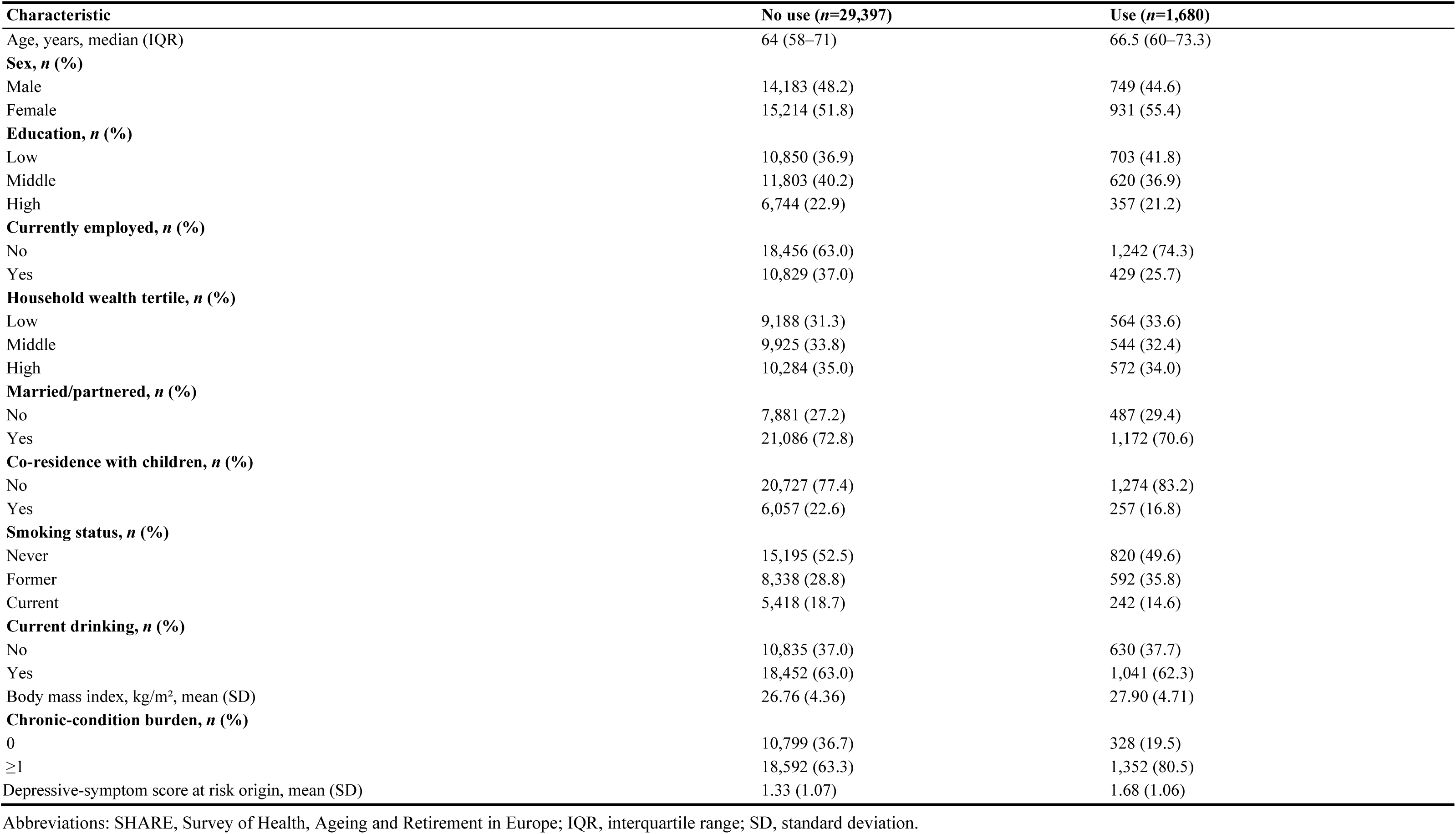
Baseline characteristics by gastrointestinal medication use in SHARE.

**Table S5.** Characteristics by two-wave gastrointestinal medication-use pattern in CHARLS.

| Characteristic | Continued non-use (*n*=3,504) | Medication initiation (*n*=227) | Medication discontinuation (*n*=192) | Use reported at both waves (*n*=164) |
| --- | --- | --- | --- | --- |
| Age, years, median (IQR) | 59 (55–65) | 60 (55–65) | 59 (55–65) | 59 (55–63) |
| **Sex, *n* (%)** |  |  |  |  |
| Male | 2,066 (59.0) | 119 (52.4) | 110 (57.3) | 99 (60.4) |
| Female | 1,438 (41.0) | 108 (47.6) | 82 (42.7) | 65 (39.6) |
| **Education, *n* (%)** |  |  |  |  |
| Low | 2,978 (85.0) | 206 (90.7) | 167 (87.0) | 141 (86.0) |
| Middle | 447 (12.8) | 16 (7.0) | 20 (10.4) | 23 (14.0) |
| High | 79 (2.3) | 5 (2.2) | 5 (2.6) | 0 (0.0) |
| **Currently employed, *n* (%)** |  |  |  |  |
| No | 1,263 (36.5) | 76 (33.6) | 59 (31.2) | 41 (25.3) |
| Yes | 2,201 (63.5) | 150 (66.4) | 130 (68.8) | 121 (74.7) |
| **Household wealth tertile, *n* (%)** |  |  |  |  |
| Low | 833 (33.0) | 56 (35.0) | 35 (25.4) | 42 (35.3) |
| Middle | 822 (32.5) | 52 (32.5) | 64 (46.4) | 44 (37.0) |
| High | 871 (34.5) | 52 (32.5) | 39 (28.3) | 33 (27.7) |
| **Married/partnered, *n* (%)** |  |  |  |  |
| No | 318 (9.1) | 16 (7.0) | 22 (11.5) | 8 (4.9) |
| Yes | 3,186 (90.9) | 211 (93.0) | 170 (88.5) | 156 (95.1) |
| **Co-residence with children, *n* (%)** |  |  |  |  |
| No | 1,567 (45.7) | 87 (38.5) | 70 (36.8) | 70 (43.5) |
| Yes | 1,861 (54.3) | 139 (61.5) | 120 (63.2) | 91 (56.5) |
| **Smoking status, *n* (%)** |  |  |  |  |
| Never | 1,903 (54.3) | 125 (55.1) | 111 (57.8) | 88 (53.7) |
| Former | 370 (10.6) | 21 (9.3) | 16 (8.3) | 20 (12.2) |
| Current | 1,229 (35.1) | 81 (35.7) | 65 (33.9) | 56 (34.1) |
| **Current drinking, *n* (%)** |  |  |  |  |
| No | 2,121 (60.5) | 149 (65.6) | 129 (67.2) | 114 (69.5) |
| Yes | 1,383 (39.5) | 78 (34.4) | 63 (32.8) | 50 (30.5) |
| Body mass index, kg/m<sup>2</sup>, mean (SD) | 23.78 (3.74) | 23.15 (3.40) | 23.04 (3.64) | 23.41 (4.05) |
| **Chronic-condition burden, *n* (%)** |  |  |  |  |
| 0 | 1,672 (48.4) | 100 (44.4) | 75 (39.9) | 54 (34.0) |
| ≥1 | 1,781 (51.6) | 125 (55.6) | 113 (60.1) | 105 (66.0) |
| Depressive-symptom score at risk origin, mean (SD) | 4.08 (2.75) | 4.52 (2.69) | 4.74 (2.59) | 4.87 (3.00) |

Abbreviations: CHARLS, China Health and Retirement Longitudinal Study; IQR, interquartile range; SD, standard deviation.

**Table S6.** Characteristics by two-wave gastrointestinal medication-use pattern in HRS.

| Characteristic | Continued non-use (*n*=8,019) | Medication initiation (*n*=482) | Medication discontinuation (*n*=390) | Use reported at both waves (*n*=910) |
| --- | --- | --- | --- | --- |
| Age, years, median (IQR) | 64 (57–73) | 67 (59–73) | 69 (60–76) | 68 (61–75) |
| **Sex, *n* (%)** |  |  |  |  |
| Male | 3,624 (45.2) | 191 (39.6) | 150 (38.5) | 346 (38.0) |
| Female | 4,395 (54.8) | 291 (60.4) | 240 (61.5) | 564 (62.0) |
| **Education, *n* (%)** |  |  |  |  |
| Low | 984 (12.3) | 83 (17.2) | 60 (15.4) | 132 (14.5) |
| Middle | 4,719 (58.9) | 307 (63.7) | 237 (60.8) | 555 (61.0) |
| High | 2,313 (28.9) | 92 (19.1) | 93 (23.8) | 223 (24.5) |
| **Currently employed, *n* (%)** |  |  |  |  |
| No | 3,955 (49.4) | 308 (63.9) | 240 (61.5) | 608 (66.8) |
| Yes | 4,059 (50.6) | 174 (36.1) | 150 (38.5) | 302 (33.2) |
| **Household wealth tertile, *n* (%)** |  |  |  |  |
| Low | 2,595 (32.4) | 193 (40.0) | 142 (36.4) | 336 (36.9) |
| Middle | 2,638 (32.9) | 148 (30.7) | 138 (35.4) | 305 (33.5) |
| High | 2,786 (34.7) | 141 (29.3) | 110 (28.2) | 269 (29.6) |
| **Married/partnered, *n* (%)** |  |  |  |  |
| No | 2,773 (34.6) | 181 (37.6) | 142 (36.4) | 333 (36.6) |
| Yes | 5,246 (65.4) | 301 (62.4) | 248 (63.6) | 577 (63.4) |
| **Co-residence with children, *n* (%)** |  |  |  |  |
| No | 5,714 (71.3) | 358 (74.3) | 302 (77.4) | 705 (77.5) |
| Yes | 2,304 (28.7) | 124 (25.7) | 88 (22.6) | 205 (22.5) |
| **Smoking status, *n* (%)** |  |  |  |  |
| Never | 3,778 (47.3) | 205 (42.8) | 164 (42.3) | 393 (43.5) |
| Former | 3,273 (41.0) | 224 (46.8) | 195 (50.3) | 432 (47.8) |
| Current | 932 (11.7) | 50 (10.4) | 29 (7.5) | 79 (8.7) |
| **Current drinking, *n* (%)** |  |  |  |  |
| No | 3,051 (38.1) | 216 (44.8) | 174 (44.6) | 427 (46.9) |
| Yes | 4,967 (61.9) | 266 (55.2) | 216 (55.4) | 483 (53.1) |
| Body mass index, kg/m<sup>2</sup>, mean (SD) | 28.35 (5.65) | 29.35 (6.10) | 29.39 (5.55) | 29.69 (6.00) |
| **Chronic-condition burden, *n* (%)** |  |  |  |  |
| 0 | 1,743 (21.7) | 35 (7.3) | 25 (6.4) | 39 (4.3) |
| ≥1 | 6,276 (78.3) | 447 (92.7) | 365 (93.6) | 871 (95.7) |
| Depressive-symptom score at risk origin, mean (SD) | 0.46 (0.66) | 0.64 (0.74) | 0.63 (0.73) | 0.61 (0.74) |

Abbreviations: HRS, Health and Retirement Study; IQR, interquartile range; SD, standard deviation.

**Table S7.** Characteristics by two-wave gastrointestinal medication-use pattern in SHARE.

| Characteristic | Continued non-use (*n*=17,142) | Medication initiation (*n*=718) | Medication discontinuation (*n*=456) | Use reported at both waves (*n*=482) |
| --- | --- | --- | --- | --- |
| Age, years, median (IQR) | 64 (58–70) | 66 (59–73) | 66 (60–73) | 67 (61–73) |
| **Sex, *n* (%)** |  |  |  |  |
| Male | 8,559 (49.9) | 319 (44.4) | 226 (49.6) | 229 (47.5) |
| Female | 8,583 (50.1) | 399 (55.6) | 230 (50.4) | 253 (52.5) |
| **Education, *n* (%)** |  |  |  |  |
| Low | 5,695 (33.2) | 298 (41.5) | 174 (38.2) | 159 (33.0) |
| Middle | 7,025 (41.0) | 273 (38.0) | 180 (39.5) | 176 (36.5) |
| High | 4,422 (25.8) | 147 (20.5) | 102 (22.4) | 147 (30.5) |
| **Currently employed, *n* (%)** |  |  |  |  |
| No | 10,186 (59.6) | 504 (70.6) | 331 (73.2) | 333 (69.2) |
| Yes | 6,910 (40.4) | 210 (29.4) | 121 (26.8) | 148 (30.8) |
| **Household wealth tertile, *n* (%)** |  |  |  |  |
| Low | 5,593 (32.6) | 263 (36.6) | 143 (31.4) | 155 (32.2) |
| Middle | 5,675 (33.1) | 226 (31.5) | 158 (34.6) | 160 (33.2) |
| High | 5,874 (34.3) | 229 (31.9) | 155 (34.0) | 167 (34.6) |
| **Married/partnered, *n* (%)** |  |  |  |  |
| No | 4,548 (26.9) | 213 (30.1) | 129 (28.5) | 142 (29.8) |
| Yes | 12,374 (73.1) | 495 (69.9) | 323 (71.5) | 335 (70.2) |
| **Co-residence with children, *n* (%)** |  |  |  |  |
| No | 12,291 (78.8) | 550 (83.3) | 338 (81.6) | 374 (86.0) |
| Yes | 3,316 (21.2) | 110 (16.7) | 76 (18.4) | 61 (14.0) |
| **Smoking status, *n* (%)** |  |  |  |  |
| Never | 8,881 (52.5) | 365 (51.5) | 219 (48.2) | 229 (48.2) |
| Former | 4,995 (29.5) | 221 (31.2) | 169 (37.2) | 189 (39.8) |
| Current | 3,041 (18.0) | 123 (17.3) | 66 (14.5) | 57 (12.0) |
| **Current drinking, *n* (%)** |  |  |  |  |
| No | 5,593 (32.7) | 281 (39.4) | 142 (31.4) | 147 (30.6) |
| Yes | 11,505 (67.3) | 433 (60.6) | 310 (68.6) | 334 (69.4) |
| Body mass index, kg/m<sup>2</sup>, mean (SD) | 26.65 (4.29) | 27.03 (4.49) | 27.75 (4.35) | 27.87 (4.62) |
| **Chronic-condition burden, *n* (%)** |  |  |  |  |
| 0 | 6,767 (39.5) | 189 (26.3) | 104 (22.8) | 105 (21.8) |
| ≥1 | 10,373 (60.5) | 529 (73.7) | 352 (77.2) | 377 (78.2) |
| Depressive-symptom score at risk origin, mean (SD) | 1.21 (1.05) | 1.30 (1.04) | 1.45 (1.06) | 1.56 (1.05) |

Abbreviations: SHARE, Survey of Health, Ageing and Retirement in Europe; IQR, interquartile range; SD, standard deviation.

**Table S8.** Sensitivity analyses for CHARLS.

| Analysis | Contrast | Primary risk-set events/N | Complete-case HR (95% CI) | Complete-case *P* | IPCW HR (95% CI) | IPCW *P* | Discrete-time HR (95% CI) | Discrete-time *P* |
| --- | --- | --- | --- | --- | --- | --- | --- | --- |
| Baseline | Gastrointestinal medication use vs no use | 2,882/6,452 | 1.22 (1.05 to 1.42) | 0.008 | 1.21 (1.07 to 1.36) | 0.002 | 1.22 (1.05 to 1.42) | 0.009 |
| Two-wave | Medication initiation | 1,464/4,087 | 1.35 (1.05 to 1.75) | 0.021 | 1.21 (0.97 to 1.51) | 0.086 | 1.37 (1.06 to 1.78) | 0.018 |
| Two-wave | Medication discontinuation | 1,464/4,087 | 1.15 (0.87 to 1.53) | 0.323 | 1.11 (0.88 to 1.40) | 0.384 | 1.13 (0.85 to 1.52) | 0.397 |
| Two-wave | Use reported at both waves | 1,464/4,087 | 1.19 (0.88 to 1.60) | 0.261 | 1.31 (1.03 to 1.66) | 0.027 | 1.23 (0.91 to 1.66) | 0.176 |

Abbreviations: HR, hazard ratio; CI, confidence interval; IPCW, inverse probability of censoring weighting.
Models were adjusted for the primary Model 4 covariate set. Complete-case and IPCW estimates were derived from Cox proportional hazards models; discrete-time estimates were derived from complementary log–log models fitted to observed survey intervals.

**Table S9.** Sensitivity analyses for HRS.

| Analysis | Contrast | Primary risk-set events/N | Complete-case HR (95% CI) | Complete-case *P* | IPCW HR (95% CI) | IPCW *P* | Discrete-time HR (95% CI) | Discrete-time *P* |
| --- | --- | --- | --- | --- | --- | --- | --- | --- |
| Baseline | Gastrointestinal medication use vs no use | 3,358/12,938 | 1.36 (1.24 to 1.48) | <0.001 | 1.35 (1.24 to 1.48) | <0.001 | 1.36 (1.25 to 1.49) | <0.001 |
| Two-wave | Medication initiation | 1,785/9,801 | 1.42 (1.17 to 1.73) | <0.001 | 1.44 (1.18 to 1.74) | <0.001 | 1.41 (1.16 to 1.71) | <0.001 |
| Two-wave | Medication discontinuation | 1,785/9,801 | 1.45 (1.17 to 1.79) | <0.001 | 1.42 (1.14 to 1.75) | 0.001 | 1.45 (1.17 to 1.79) | <0.001 |
| Two-wave | Use reported at both waves | 1,785/9,801 | 1.30 (1.12 to 1.50) | <0.001 | 1.29 (1.11 to 1.49) | <0.001 | 1.30 (1.12 to 1.50) | <0.001 |

Abbreviations: HR, hazard ratio; CI, confidence interval; IPCW, inverse probability of censoring weighting.
Models were adjusted for the primary Model 4 covariate set. Complete-case and IPCW estimates were derived from Cox proportional hazards models; discrete-time estimates were derived from complementary log–log models fitted to observed survey intervals.

**Table S10.**
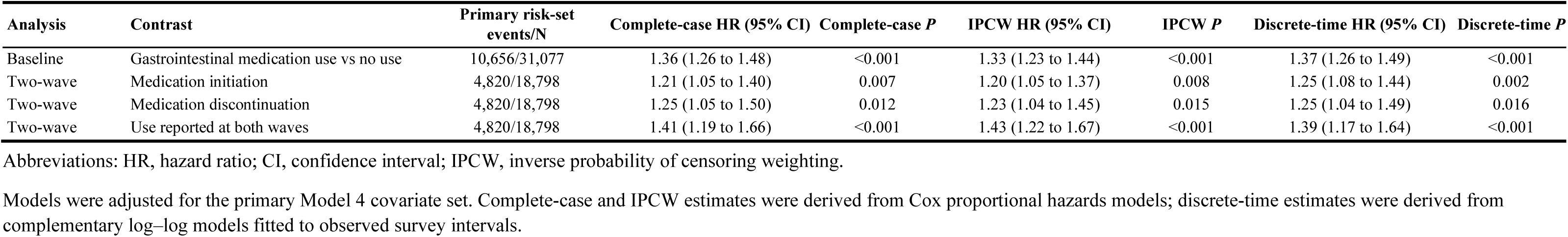
Sensitivity analyses for SHARE.

| Analysis | Contrast | Primary risk-set events/N | Complete-case HR (95% CI) | Complete-case *P* | IPCW HR (95% CI) | IPCW *P* | Discrete-time HR (95% CI) | Discrete-time *P* |
| --- | --- | --- | --- | --- | --- | --- | --- | --- |
| Baseline | Gastrointestinal medication use vs no use | 10,656/31,077 | 1.36 (1.26 to 1.48) | <0.001 | 1.33 (1.23 to 1.44) | <0.001 | 1.37 (1.26 to 1.49) | <0.001 |
| Two-wave | Medication initiation | 4,820/18,798 | 1.21 (1.05 to 1.40) | 0.007 | 1.20 (1.05 to 1.37) | 0.008 | 1.25 (1.08 to 1.44) | 0.002 |
| Two-wave | Medication discontinuation | 4,820/18,798 | 1.25 (1.05 to 1.50) | 0.012 | 1.23 (1.04 to 1.45) | 0.015 | 1.25 (1.04 to 1.49) | 0.016 |
| Two-wave | Use reported at both waves | 4,820/18,798 | 1.41 (1.19 to 1.66) | <0.001 | 1.43 (1.22 to 1.67) | <0.001 | 1.39 (1.17 to 1.64) | <0.001 |

Abbreviations: HR, hazard ratio; CI, confidence interval; IPCW, inverse probability of censoring weighting.
Models were adjusted for the primary Model 4 covariate set. Complete-case and IPCW estimates were derived from Cox proportional hazards models; discrete-time estimates were derived from complementary log–log models fitted to observed survey intervals.

**Table S11.** Rolling person-period sensitivity analysis.

| **Cohort** | **Contrast** | **Reference** | **HR (95% CI)** | ***P* value** | **Participants** | **Person-periods** | **Events** |
| --- | --- | --- | --- | --- | --- | --- | --- |
| CHARLS | Medication initiation | Continued non-use | 1.11 (0.87 to 1.43) | 0.396 | 3,702 | 7,577 | 1,226 |
| CHARLS | Medication discontinuation | Continued non-use | 1.32 (1.02 to 1.71) | 0.032 | 3,702 | 7,577 | 1,226 |
| CHARLS | Use reported at both waves | Continued non-use | 1.29 (1.00 to 1.65) | 0.046 | 3,702 | 7,577 | 1,226 |
| HRS | Medication initiation | Continued non-use | 1.56 (1.29 to 1.87) | <0.001 | 9,520 | 22,390 | 1,691 |
| HRS | Medication discontinuation | Continued non-use | 1.30 (1.05 to 1.60) | 0.014 | 9,520 | 22,390 | 1,691 |
| HRS | Use reported at both waves | Continued non-use | 1.35 (1.16 to 1.56) | <0.001 | 9,520 | 22,390 | 1,691 |
| SHARE | Medication initiation | Continued non-use | 1.28 (1.09 to 1.50) | 0.003 | 16,843 | 27,129 | 3,099 |
| SHARE | Medication discontinuation | Continued non-use | 1.21 (1.00 to 1.47) | 0.053 | 16,843 | 27,129 | 3,099 |
| SHARE | Use reported at both waves | Continued non-use | 1.41 (1.18 to 1.68) | <0.001 | 16,843 | 27,129 | 3,099 |

Abbreviations: CHARLS, China Health and Retirement Longitudinal Study; HRS, Health and Retirement Study; SHARE, Survey of Health, Ageing and Retirement in Europe; HR, hazard ratio; CI, confidence interval.

**Table S12.** Cross-cohort pooled estimates of gastrointestinal medication use and incident depressive symptoms, by model.

| Contrast | Model | Pooled HR (95% CI) | $I^2$ (%) | $\tau^2$ | $P$ for heterogeneity | 95% prediction interval | E-value (CI limit) |
| --- | --- | --- | --- | --- | --- | --- | --- |
| Baseline use vs no use | Model 1 | 1.45 (1.26–1.67) | 69 | 0.0133 | 0.039 | 1.11–1.89 | 1.91 (1.62) |
| Baseline use vs no use | Model 2 | 1.41 (1.27–1.57) | 54 | 0.0068 | 0.112 | 1.16–1.71 | 1.85 (1.64) |
| Baseline use vs no use | Model 3 | 1.37 (1.27–1.48) | 32 | 0.0024 | 0.230 | 1.21–1.55 | 1.79 (1.64) |
| Baseline use vs no use | Model 4 | 1.31 (1.21–1.41) | 29 | 0.0020 | 0.244 | 1.16–1.47 | 1.70 (1.55) |
| Medication discontinuation | Model 1 | 1.37 (1.11–1.71) | 51 | 0.0262 | 0.128 | 0.94–2.02 | 1.80 (1.35) |
| Medication discontinuation | Model 2 | 1.31 (1.10–1.56) | 36 | 0.0134 | 0.211 | 0.98–1.75 | 1.71 (1.34) |
| Medication discontinuation | Model 3 | 1.29 (1.12–1.49) | 22 | 0.0050 | 0.280 | 1.06–1.58 | 1.68 (1.39) |
| Medication discontinuation | Model 4 | 1.25 (1.11–1.40) | 12 | 0.0000 | 0.319 | 1.11–1.40 | 1.60 (1.36) |
| Medication initiation | Model 1 | 1.44 (1.21–1.70) | 41 | 0.0142 | 0.184 | 1.08–1.92 | 1.89 (1.55) |
| Medication initiation | Model 2 | 1.36 (1.13–1.64) | 50 | 0.0180 | 0.135 | 0.99–1.88 | 1.78 (1.40) |
| Medication initiation | Model 3 | 1.30 (1.14–1.49) | 20 | 0.0057 | 0.285 | 1.07–1.59 | 1.69 (1.42) |
| Medication initiation | Model 4 | 1.27 (1.13–1.43) | 8 | 0.0032 | 0.337 | 1.08–1.49 | 1.64 (1.39) |
| Use reported at both waves | Model 1 | 1.50 (1.36–1.65) | 0 | 0.0000 | 0.639 | 1.36–1.65 | 1.98 (1.78) |
| Use reported at both waves | Model 2 | 1.45 (1.32–1.59) | 0 | 0.0000 | 0.842 | 1.32–1.59 | 1.91 (1.71) |
| Use reported at both waves | Model 3 | 1.41 (1.28–1.55) | 0 | 0.0000 | 0.603 | 1.28–1.55 | 1.85 (1.65) |
| Use reported at both waves | Model 4 | 1.34 (1.22–1.47) | 0 | 0.0000 | 0.652 | 1.22–1.47 | 1.75 (1.55) |

Abbreviations: CHARLS, China Health and Retirement Longitudinal Study; HRS, Health and Retirement Study; SHARE, Survey of Health, Ageing and Retirement in Europe; CI, confidence interval; HR, hazard ratio; REML, restricted maximum likelihood; $I^2$ , percentage of variability attributable to between-cohort heterogeneity; $\tau^2$ , between-cohort variance.
Pooled estimates are from inverse-variance weighted random-effects models with $\tau^2$ estimated by restricted maximum likelihood. $I^2$ is the percentage of variability attributable to between-cohort heterogeneity; the 95% prediction interval indicates the range of estimates expected in a future cohort. E-values were derived after converting hazard ratios to approximate risk ratios. $I^2$ was derived from Cochran's $Q$ statistic, whereas $\tau^2$ was estimated by restricted maximum likelihood and truncated at zero; when $\tau^2$ is zero the prediction interval coincides with the confidence interval.

**Table S13.** Stratum-specific associations of baseline gastrointestinal medication use with incident depressive symptoms, and tests for effect modification.

| Cohort | Domain | Subgroup variable | Level | n / events | HR (95% CI) | P for interaction |
| --- | --- | --- | --- | --- | --- | --- |
| CHARLS | Demographic | Age, years | 50–64 | 4,592/2,115 | 1.24 (1.08–1.42) | 0.252 |
| CHARLS | Demographic | Age, years | 65–74 | 1,445/611 | 1.03 (0.79–1.34) | 0.252 |
| CHARLS | Demographic | Age, years | ≥75 | 415/156 | 1.70 (0.91–3.17) | 0.252 |
| CHARLS | Demographic | Sex | Men | 3,590/1,410 | 1.11 (0.93–1.33) | 0.236 |
| CHARLS | Demographic | Sex | Women | 2,862/1,472 | 1.29 (1.10–1.52) | 0.236 |
| CHARLS | Socioeconomic | Education | Low | 5,552/2,614 | 1.23 (1.08–1.39) | 0.310 |
| CHARLS | Socioeconomic | Education | Middle/High | 900/268 | 0.96 (0.61–1.51) | 0.310 |
| CHARLS | Socioeconomic | Current employment | Not working | 2,478/1,010 | 1.09 (0.87–1.36) | 0.272 |
| CHARLS | Socioeconomic | Current employment | Working | 3,974/1,872 | 1.26 (1.09–1.45) | 0.272 |
| CHARLS | Socioeconomic | Household wealth tertile | Low | 2,039/1,008 | 1.28 (1.03–1.59) | 0.617 |
| CHARLS | Socioeconomic | Household wealth tertile | Middle | 2,204/1,044 | 1.11 (0.90–1.36) | 0.617 |
| CHARLS | Socioeconomic | Household wealth tertile | High | 2,214/830 | 1.26 (0.99–1.60) | 0.617 |
| CHARLS | Household | Marital/partner status | Not partnered | 681/311 | 1.02 (0.66–1.56) | 0.411 |
| CHARLS | Household | Marital/partner status | Partnered | 5,771/2,571 | 1.23 (1.08–1.39) | 0.411 |
| CHARLS | Household | Living with children | No | 2,844/1,229 | 1.21 (1.00–1.45) | 0.976 |
| CHARLS | Household | Living with children | Yes | 3,608/1,653 | 1.20 (1.03–1.41) | 0.976 |
| CHARLS | Lifestyle | Smoking status | Never | 3,616/1,724 | 1.23 (1.06–1.43) | 0.733 |
| CHARLS | Lifestyle | Smoking status | Former | 649/244 | 1.33 (0.88–2.00) | 0.733 |
| CHARLS | Lifestyle | Smoking status | Current | 2,187/914 | 1.13 (0.91–1.41) | 0.733 |
| CHARLS | Lifestyle | Current/recent drinking | No | 4,113/1,947 | 1.19 (1.03–1.37) | 0.708 |
| CHARLS | Lifestyle | Current/recent drinking | Yes | 2,339/935 | 1.25 (1.00–1.57) | 0.708 |
| CHARLS | Health burden | Number of chronic conditions | 0 | 2,880/1,188 | 1.19 (0.97–1.46) | 0.883 |
| CHARLS | Health burden | Number of chronic conditions | ≥1 | 3,572/1,694 | 1.21 (1.05–1.40) | 0.883 |
| HRS | Demographic | Age, years | 50–64 | 6,170/1,606 | 1.39 (1.21–1.60) | 0.779 |
| HRS | Demographic | Age, years | 65–74 | 3,608/907 | 1.38 (1.18–1.61) | 0.779 |
| HRS | Demographic | Age, years | ≥75 | 3,160/845 | 1.29 (1.09–1.53) | 0.779 |
| HRS | Demographic | Sex | Men | 5,586/1,213 | 1.32 (1.13–1.54) | 0.710 |
| HRS | Demographic | Sex | Women | 7,352/2,145 | 1.37 (1.23–1.52) | 0.710 |
| HRS | Socioeconomic | Education | Low | 1,893/643 | 1.66 (1.38–2.00) | 0.015 |
| HRS | Socioeconomic | Education | Middle/High | 11,045/2,715 | 1.28 (1.16–1.41) | 0.015 |
| HRS | Socioeconomic | Current employment | Not working | 7,139/2,050 | 1.35 (1.22–1.50) | 0.991 |
| HRS | Socioeconomic | Current employment | Working | 5,799/1,308 | 1.35 (1.15–1.58) | 0.991 |
| HRS | Socioeconomic | Household wealth tertile | Low | 4,286/1,411 | 1.46 (1.28–1.66) | 0.253 |
| HRS | Socioeconomic | Household wealth tertile | Middle | 4,316/1,054 | 1.32 (1.13–1.55) | 0.253 |
| HRS | Socioeconomic | Household wealth tertile | High | 4,336/893 | 1.21 (1.00–1.46) | 0.253 |
| HRS | Household | Marital/partner status | Not partnered | 4,801/1,402 | 1.35 (1.18–1.54) | 0.964 |
| HRS | Household | Marital/partner status | Partnered | 8,137/1,956 | 1.36 (1.21–1.52) | 0.964 |
| HRS | Household | Living with children | No | 9,395/2,434 | 1.35 (1.22–1.50) | 0.977 |
| HRS | Household | Living with children | Yes | 3,543/924 | 1.35 (1.13–1.61) | 0.977 |
| HRS | Lifestyle | Smoking status | Never | 5,919/1,415 | 1.23 (1.07–1.42) | 0.222 |
| HRS | Lifestyle | Smoking status | Former | 5,432/1,384 | 1.45 (1.28–1.64) | 0.222 |
| HRS | Lifestyle | Smoking status | Current | 1,588/558 | 1.41 (1.12–1.79) | 0.222 |
| HRS | Lifestyle | Current/recent drinking | No | 5,379/1,525 | 1.31 (1.15–1.48) | 0.438 |
| HRS | Lifestyle | Current/recent drinking | Yes | 7,559/1,833 | 1.40 (1.24–1.58) | 0.438 |
| HRS | Health burden | Number of chronic conditions | 0 | 2,232/381 | 1.47 (0.93–2.33) | 0.719 |
| HRS | Health burden | Number of chronic conditions | ≥1 | 10,706/2,977 | 1.35 (1.23–1.48) | 0.719 |
| SHARE | Demographic | Age, years | 50–64 | 16,297/4,913 | 1.42 (1.25–1.60) | 0.255 |
| SHARE | Demographic | Age, years | 65–74 | 9,534/3,490 | 1.37 (1.21–1.55) | 0.255 |
| SHARE | Demographic | Age, years | ≥75 | 5,246/2,253 | 1.21 (1.05–1.40) | 0.255 |

| Cohort | Domain | Subgroup variable | Level | *n* / events | HR (95% CI) | *P* for interaction |
| --- | --- | --- | --- | --- | --- | --- |
| SHARE | Demographic | Sex | Men | 14,932/4,119 | 1.31 (1.16–1.48) | 0.669 |
| SHARE | Demographic | Sex | Women | 16,145/6,537 | 1.35 (1.23–1.49) | 0.669 |
| SHARE | Socioeconomic | Education | Low | 11,553/4,710 | 1.36 (1.22–1.52) | 0.640 |
| SHARE | Socioeconomic | Education | Middle/High | 19,524/5,946 | 1.31 (1.18–1.46) | 0.640 |
| SHARE | Socioeconomic | Current employment | Not working | 19,802/7,615 | 1.34 (1.23–1.46) | 0.883 |
| SHARE | Socioeconomic | Current employment | Working | 11,275/3,041 | 1.32 (1.12–1.57) | 0.883 |
| SHARE | Socioeconomic | Household wealth tertile | Low | 9,752/3,822 | 1.30 (1.15–1.48) | 0.837 |
| SHARE | Socioeconomic | Household wealth tertile | Middle | 10,469/3,657 | 1.34 (1.18–1.53) | 0.837 |
| SHARE | Socioeconomic | Household wealth tertile | High | 10,856/3,177 | 1.38 (1.20–1.57) | 0.837 |
| SHARE | Household | Marital/partner status | Not partnered | 8,466/3,116 | 1.33 (1.16–1.52) | 0.895 |
| SHARE | Household | Marital/partner status | Partnered | 22,612/7,540 | 1.34 (1.23–1.47) | 0.895 |
| SHARE | Household | Living with children | No | 24,147/8,418 | 1.34 (1.23–1.45) | 0.957 |
| SHARE | Household | Living with children | Yes | 6,930/2,238 | 1.34 (1.11–1.63) | 0.957 |
| SHARE | Lifestyle | Smoking status | Never | 16,241/5,802 | 1.36 (1.22–1.51) | 0.885 |
| SHARE | Lifestyle | Smoking status | Former | 9,074/2,889 | 1.30 (1.14–1.48) | 0.885 |
| SHARE | Lifestyle | Smoking status | Current | 5,765/1,967 | 1.35 (1.11–1.66) | 0.885 |
| SHARE | Lifestyle | Current/recent drinking | No | 11,528/4,728 | 1.31 (1.17–1.47) | 0.638 |
| SHARE | Lifestyle | Current/recent drinking | Yes | 19,550/5,928 | 1.36 (1.23–1.50) | 0.638 |
| SHARE | Health burden | Number of chronic conditions | 0 | 11,128/2,945 | 1.31 (1.08–1.59) | 0.820 |
| SHARE | Health burden | Number of chronic conditions | ≥1 | 19,949/7,711 | 1.34 (1.24–1.46) | 0.820 |

Abbreviations: CHARLS, China Health and Retirement Longitudinal Study; HRS, Health and Retirement Study; SHARE, Survey of Health, Ageing and Retirement in Europe; CI, confidence interval; HR, hazard ratio.
Model 4 adjustment within each cohort. *n* / events are median numbers of participants and events across the multiply imputed datasets. *P* for interaction is from the exposure-by-subgroup product term. Subgroup *n* are medians across the 20 imputed datasets and are rounded, so stratum counts may not sum exactly to the cohort total.

**Table S14.**
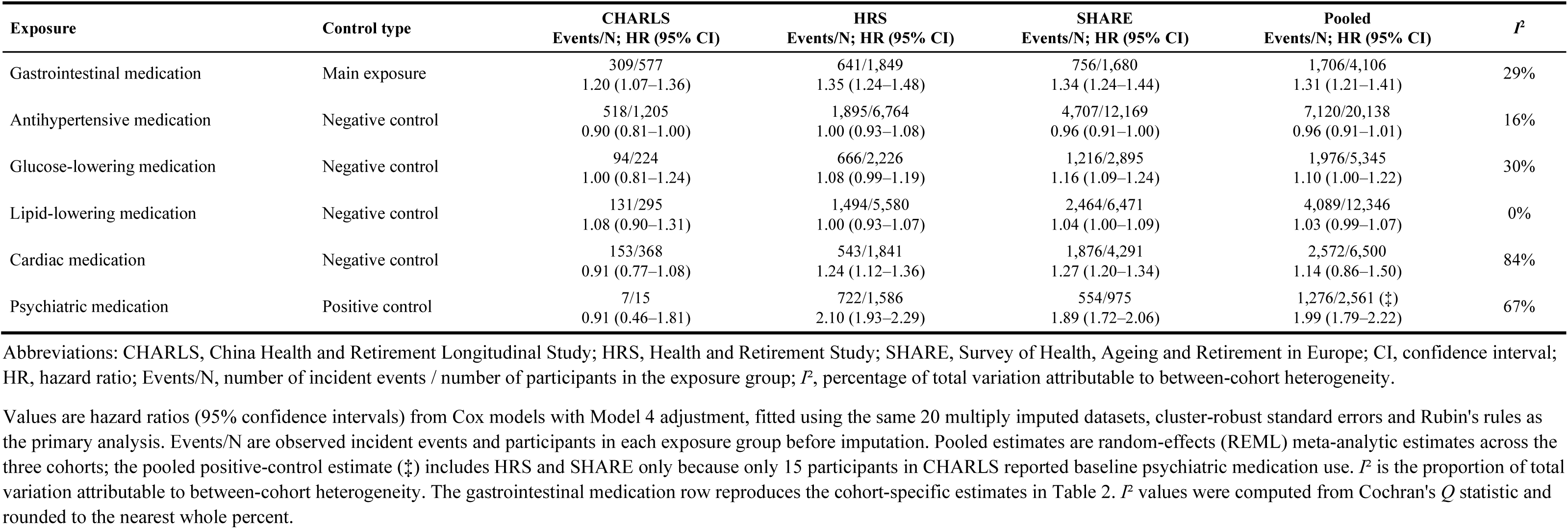
Negative- and positive-control exposures and incident depressive symptoms, with observed events/N and pooled estimates, by cohort.

**Table S15.**
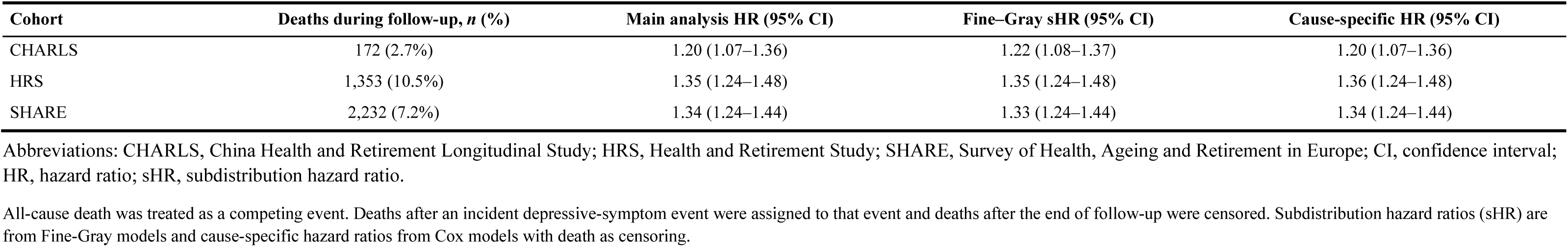
Associations of baseline gastrointestinal medication use with incident depressive symptoms accounting for death as a competing event.

| Cohort | Deaths during follow-up, *n* (%) | Main analysis HR (95% CI) | Fine-Gray sHR (95% CI) | Cause-specific HR (95% CI) |
| --- | --- | --- | --- | --- |
| CHARLS | 172 (2.7%) | 1.20 (1.07–1.36) | 1.22 (1.08–1.37) | 1.20 (1.07–1.36) |
| HRS | 1,353 (10.5%) | 1.35 (1.24–1.48) | 1.35 (1.24–1.48) | 1.36 (1.24–1.48) |
| SHARE | 2,232 (7.2%) | 1.34 (1.24–1.44) | 1.33 (1.24–1.44) | 1.34 (1.24–1.44) |

Abbreviations: CHARLS, China Health and Retirement Longitudinal Study; HRS, Health and Retirement Study; SHARE, Survey of Health, Ageing and Retirement in Europe; CI, confidence interval; HR, hazard ratio; sHR, subdistribution hazard ratio.
All-cause death was treated as a competing event. Deaths after an incident depressive-symptom event were assigned to that event and deaths after the end of follow-up were censored. Subdistribution hazard ratios (sHR) are from Fine-Gray models and cause-specific hazard ratios from Cox models with death as censoring.

**Table S16.** Sensitivity analyses excluding participants who reported psychiatric medication use or mental illness at baseline.

| Cohort | Analytic sample | *n* | HR (95% CI) | *P* |
| --- | --- | --- | --- | --- |
| CHARLS | Main analysis (eight-condition burden) | 6,452 | 1.20 (1.07 to 1.36) | 0.002 |
| CHARLS | Main analysis, seven physical conditions only | 6,452 | 1.21 (1.07 to 1.36) | 0.002 |
| CHARLS | Excluding psychiatric medication users | 6,437 | 1.21 (1.07 to 1.36) | 0.002 |
| CHARLS | Excluding psychiatric medication users and mental illness | 6,415 | 1.21 (1.07 to 1.36) | 0.002 |
| HRS | Main analysis (eight-condition burden) | 12,938 | 1.35 (1.24 to 1.48) | < 0.001 |
| HRS | Main analysis, seven physical conditions only | 12,938 | 1.36 (1.24 to 1.48) | < 0.001 |
| HRS | Excluding psychiatric medication users | 11,332 | 1.26 (1.13 to 1.41) | < 0.001 |
| HRS | Excluding psychiatric medication users and mental illness | 10,752 | 1.26 (1.12 to 1.41) | < 0.001 |
| SHARE | Main analysis (eight-condition burden) | 31,077 | 1.34 (1.24 to 1.44) | < 0.001 |
| SHARE | Main analysis, seven physical conditions only | 31,077 | 1.34 (1.25 to 1.45) | < 0.001 |
| SHARE | Excluding psychiatric medication users | 30,102 | 1.32 (1.22 to 1.43) | < 0.001 |
| SHARE | Excluding psychiatric medication users and mental illness | 28,894 | 1.33 (1.22 to 1.44) | < 0.001 |

All models use the Model 4 covariate set and the same 20 multiply imputed datasets as the main analysis, with estimates combined using Rubin's rules. The chronic-condition burden excludes mental illness in all scenarios except the first, which reproduces the main analysis. Abbreviations: CHARLS, China Health and Retirement Longitudinal Study; HRS, Health and Retirement Study; SHARE, Survey of Health, Ageing and Retirement in Europe; CI, confidence interval; HR, hazard ratio.

**Table S17.** Sensitivity analyses with additional adjustment for the baseline depressive-symptom score or with chronic-condition burden categorise.

| Cohort | Model | HR (95% CI) | P |
| --- | --- | --- | --- |
| CHARLS | Main model (Model 4) | 1.20 (1.07 to 1.36) | 0.002 |
| CHARLS | Additional adjustment for baseline symptom score | 1.12 (1.00 to 1.27) | 0.055 |
| CHARLS | Chronic-condition burden (0/1/2/≥3) | 1.19 (1.06 to 1.34) | 0.004 |
| HRS | Main model (Model 4) | 1.35 (1.24 to 1.48) | < 0.001 |
| HRS | Additional adjustment for baseline symptom score | 1.23 (1.12 to 1.34) | < 0.001 |
| HRS | Chronic-condition burden (0/1/2/≥3) | 1.24 (1.14 to 1.36) | < 0.001 |
| SHARE | Main model (Model 4) | 1.34 (1.24 to 1.44) | < 0.001 |
| SHARE | Additional adjustment for baseline symptom score | 1.23 (1.14 to 1.33) | < 0.001 |
| SHARE | Chronic-condition burden (0/1/2/≥3) | 1.26 (1.17 to 1.36) | < 0.001 |

All models use the Model 4 covariate set and the same 20 multiply imputed datasets, with estimates combined using Rubin's rules. Abbreviations: CHARLS, China Health and Retirement Longitudinal Study; HRS, Health and Retirement Study; SHARE, Survey of Health, Ageing and Retirement in Europe; CI, confidence interval; HR, hazard ratio.

**Table S18.** Associations of digestive-disease diagnosis and gastrointestinal medication use with incident depressive symptoms.

| Cohort | Group | HR (95% CI) | P |
| --- | --- | --- | --- |
| CHARLS | Digestive disease, no medication (*n*=646; 320 events) | 1.00 (reference) | — |
| CHARLS | No digestive disease, no medication (*n*=5,229; 2,253 events) | 0.87 (0.77 to 0.98) | 0.024 |
| CHARLS | Digestive disease, medication (*n*=434; 242 events) | 1.14 (0.96 to 1.35) | 0.133 |
| CHARLS | No digestive disease, medication (*n*=143; 67 events) | 0.87 (0.66 to 1.14) | 0.316 |
| SHARE | Digestive disease, no medication (*n*=1,337; 560 events) | 1.00 (reference) | — |
| SHARE | No digestive disease, no medication (*n*=28,058; 9,338 events) | 0.82 (0.75 to 0.89) | < 0.001 |
| SHARE | Digestive disease, medication (*n*=459; 227 events) | 1.21 (1.03 to 1.42) | 0.018 |
| SHARE | No digestive disease, medication (*n*=1,221; 529 events) | 1.07 (0.94 to 1.20) | 0.302 |
| Pooled (CHARLS + SHARE) | Disease, medication vs disease, no medication | 1.18 (1.05 to 1.32) | 0.006 |

Model 4 covariate set and the same 20 multiply imputed datasets; estimates combined using Rubin's rules. HRS is not shown because the harmonised dataset does not include a self-reported digestive-disease diagnosis. In CHARLS, the ever-diagnosed digestive-disease indicator and current-medication item were harmonised from separate survey questions across waves, resulting in 143 participants with recorded medication use without a concurrent harmonised diagnosis. Abbreviations: CHARLS, China Health and Retirement Longitudinal Study; SHARE, Survey of Health, Ageing and Retirement in Europe; CI, confidence interval; HR, hazard ratio. The pooled estimate combines CHARLS and SHARE using random-effects meta-analysis with REML; *P* = 0%.

**Table S19.** Sensitivity analyses of the two-wave medication-use patterns.

| **Cohort** | **Analytic sample** | **Pattern** | **HR (95% CI)** | **P** |
| --- | --- | --- | --- | --- |
| CHARLS | Additional baseline-score adjustment | Initiation | 1.18 (0.94 to 1.47) | 0.147 |
| CHARLS | Additional baseline-score adjustment | Discontinuation | 1.03 (0.81 to 1.30) | 0.810 |
| CHARLS | Additional baseline-score adjustment | Use at both waves | 1.20 (0.95 to 1.52) | 0.130 |
| CHARLS | Excluding psychiatric medication users | Initiation | 1.19 (0.95 to 1.48) | 0.130 |
| CHARLS | Excluding psychiatric medication users | Discontinuation | 1.10 (0.87 to 1.39) | 0.420 |
| CHARLS | Excluding psychiatric medication users | Use at both waves | 1.31 (1.03 to 1.65) | 0.026 |
| HRS | Additional baseline-score adjustment | Initiation | 1.31 (1.08 to 1.59) | 0.006 |
| HRS | Additional baseline-score adjustment | Discontinuation | 1.29 (1.04 to 1.60) | 0.019 |
| HRS | Additional baseline-score adjustment | Use at both waves | 1.20 (1.04 to 1.39) | 0.014 |
| HRS | Excluding psychiatric medication users | Initiation | 1.53 (1.24 to 1.89) | < 0.001 |
| HRS | Excluding psychiatric medication users | Discontinuation | 1.55 (1.22 to 1.97) | < 0.001 |
| HRS | Excluding psychiatric medication users | Use at both waves | 1.18 (0.99 to 1.41) | 0.069 |
| SHARE | Additional baseline-score adjustment | Initiation | 1.19 (1.04 to 1.36) | 0.011 |
| SHARE | Additional baseline-score adjustment | Discontinuation | 1.15 (0.97 to 1.36) | 0.106 |
| SHARE | Additional baseline-score adjustment | Use at both waves | 1.30 (1.11 to 1.53) | 0.001 |
| SHARE | Excluding psychiatric medication users | Initiation | 1.18 (1.03 to 1.35) | 0.020 |
| SHARE | Excluding psychiatric medication users | Discontinuation | 1.21 (1.02 to 1.44) | 0.029 |
| SHARE | Excluding psychiatric medication users | Use at both waves | 1.39 (1.18 to 1.64) | < 0.001 |

Model 4 covariate set and the same 20 multiply imputed datasets; estimates combined using Rubin's rules. Continued non-use is the reference category in every model. Abbreviations: CHARLS, China Health and Retirement Longitudinal Study; HRS, Health and Retirement Study; SHARE, Survey of Health, Ageing and Retirement in Europe; CI, confidence interval; HR, hazard ratio.

**Table S20.** Sensitivity analysis with age modelled by a natural cubic spline.

| Analysis and exposure contrast | CHARLS HR (95% CI) | HRS HR (95% CI) | SHARE HR (95% CI) |
| --- | --- | --- | --- |
| **Baseline analysis** |  |  |  |
| No use | 1.00 (Reference) | 1.00 (Reference) | 1.00 (Reference) |
| Gastrointestinal medication use | 1.22 (1.09 to 1.38) | 1.36 (1.25 to 1.49) | 1.34 (1.24 to 1.45) |
| **Two-wave pattern analysis** |  |  |  |
| Continued non-use | 1.00 (Reference) | 1.00 (Reference) | 1.00 (Reference) |
| Medication initiation | 1.21 (0.97 to 1.50) | 1.44 (1.19 to 1.75) | 1.21 (1.06 to 1.38) |
| Medication discontinuation | 1.11 (0.88 to 1.40) | 1.41 (1.14 to 1.75) | 1.23 (1.04 to 1.45) |
| Use reported at both waves | 1.30 (1.03 to 1.65) | 1.31 (1.13 to 1.51) | 1.44 (1.23 to 1.68) |

All models used the Model 4 covariate set across the 20 multiply imputed datasets, with estimates combined using Rubin's rules. Linear age was replaced by a natural cubic spline with knots placed at the cohort-specific tertiles of observed baseline age. Abbreviations: CHARLS, China Health and Retirement Longitudinal Study; CI, confidence interval; HR, hazard ratio; HRS, Health and Retirement Study; SHARE, Survey of Health, Ageing and Retirement in Europe.

**Table S21.** Participant counts and incident depressive-symptom events by SHARE country.

| Country | Baseline analysis: participants (*n*) | Baseline analysis: incident events (*n*) | Two-wave analysis: participants (*n*) | Two-wave analysis: incident events (*n*) |
| --- | --- | --- | --- | --- |
| Austria | 3,125 | 877 | 2,070 | 469 |
| Belgium | 2,895 | 976 | 1,956 | 503 |
| Czech Republic | 3,129 | 1,049 | 1,988 | 462 |
| Denmark | 1,613 | 418 | 1,267 | 250 |
| Estonia | 3,306 | 1,420 | 2,004 | 607 |
| France | 1,865 | 352 | 983 | 179 |
| Germany | 828 | 287 | 643 | 171 |
| Hungary | 769 | 201 | — | — |
| Italy | 1,871 | 910 | 1,176 | 419 |
| Netherlands | 2,454 | 662 | 1,842 | 377 |
| Poland | 788 | 374 | — | — |
| Portugal | 852 | 344 | — | — |
| Slovenia | 1,599 | 566 | 1,027 | 266 |
| Spain | 1,962 | 754 | 1,327 | 408 |
| Sweden | 1,302 | 404 | 913 | 231 |
| Switzerland | 2,719 | 1,062 | 1,602 | 478 |
| **Total** | **31,077** | **10,656** | **18,798** | **4,820** |

Poland, Hungary, and Portugal were excluded from the two-wave change analysis because they lacked a comparable second-round medication assessment at Wave 5 (shown as em dashes). Country identifiers follow the SHARE numeric code scheme (Switzerland aggregates the German, French, and Italian language regions; Belgium aggregates the French and Flemish regions). Abbreviations: *n*, number of participants; events, incident depressive-symptom events over follow-up.

**Table S22.** Missing data rates for covariates in the baseline analytic sample.

| Covariate | CHARLS (*n*=6,452) | HRS (*n*=12,938) | SHARE (*n*=31,077) |
| --- | --- | --- | --- |
| Age, years | 0 (0.0) | 0 (0.0) | 0 (0.0) |
| Female sex | 0 (0.0) | 0 (0.0) | 0 (0.0) |
| Education | 0 (0.0) | 3 (0.0) | 0 (0.0) |
| Currently employed | 86 (1.3) | 10 (0.1) | 121 (0.4) |
| Household wealth tertile | 1,844 (28.6) | 0 (0.0) | 0 (0.0) |
| Married/partnered | 0 (0.0) | 0 (0.0) | 451 (1.5) |
| Co-residing with at least one child | 141 (2.2) | 1 (0.0) | 2,762 (8.9) |
| Smoking status | 3 (0.0) | 66 (0.5) | 472 (1.5) |
| Current/recent alcohol use | 0 (0.0) | 3 (0.0) | 119 (0.4) |
| BMI, kg/m<sup>2</sup> | 928 (14.4) | 152 (1.2) | 912 (2.9) |
| Chronic-condition burden | 0 (0.0) | 0 (0.0) | 0 (0.0) |

Values are number (percentage) of participants with missing values among the baseline analytic sample (CHARLS *n*=6,452; HRS *n*=12,938; SHARE *n*=31,077). Missing covariates were imputed by multiple imputation by chained equations (20 imputed datasets; estimates combined by Rubin's rules). Age, sex, education and chronic-condition burden were complete in all three cohorts.

**Figure S1.**
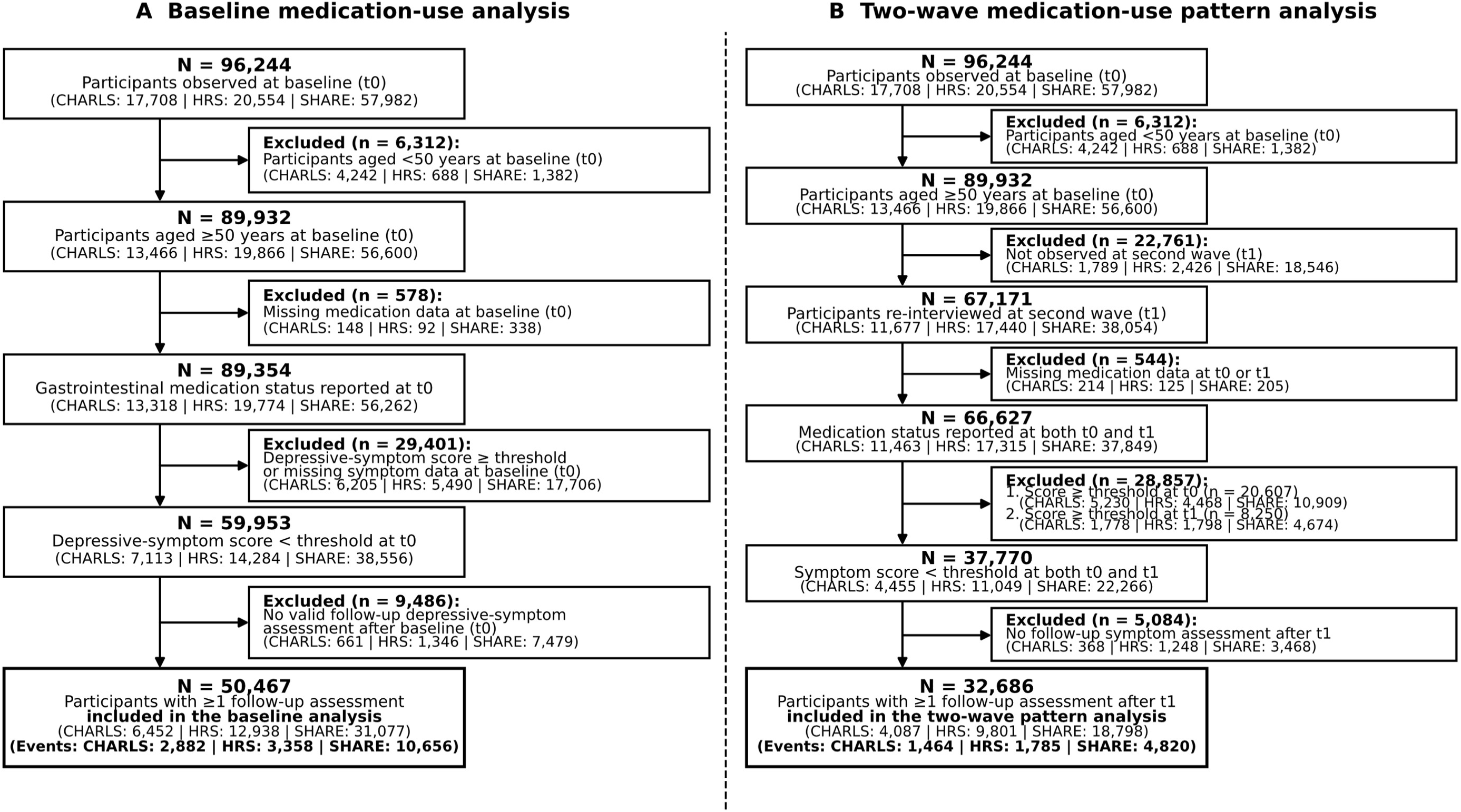
Cohort-specific participant flow for the baseline and two-wave analyses, shown separately for CHARLS, HRS and SHARE. Numbers are participants remaining at each step (from the observed survey data); the final box gives the analytic sample and the number of incident depressive-symptom events. Further exclusion reasons and combined totals are summarised in Fig. 1B.

**Figure S2.**
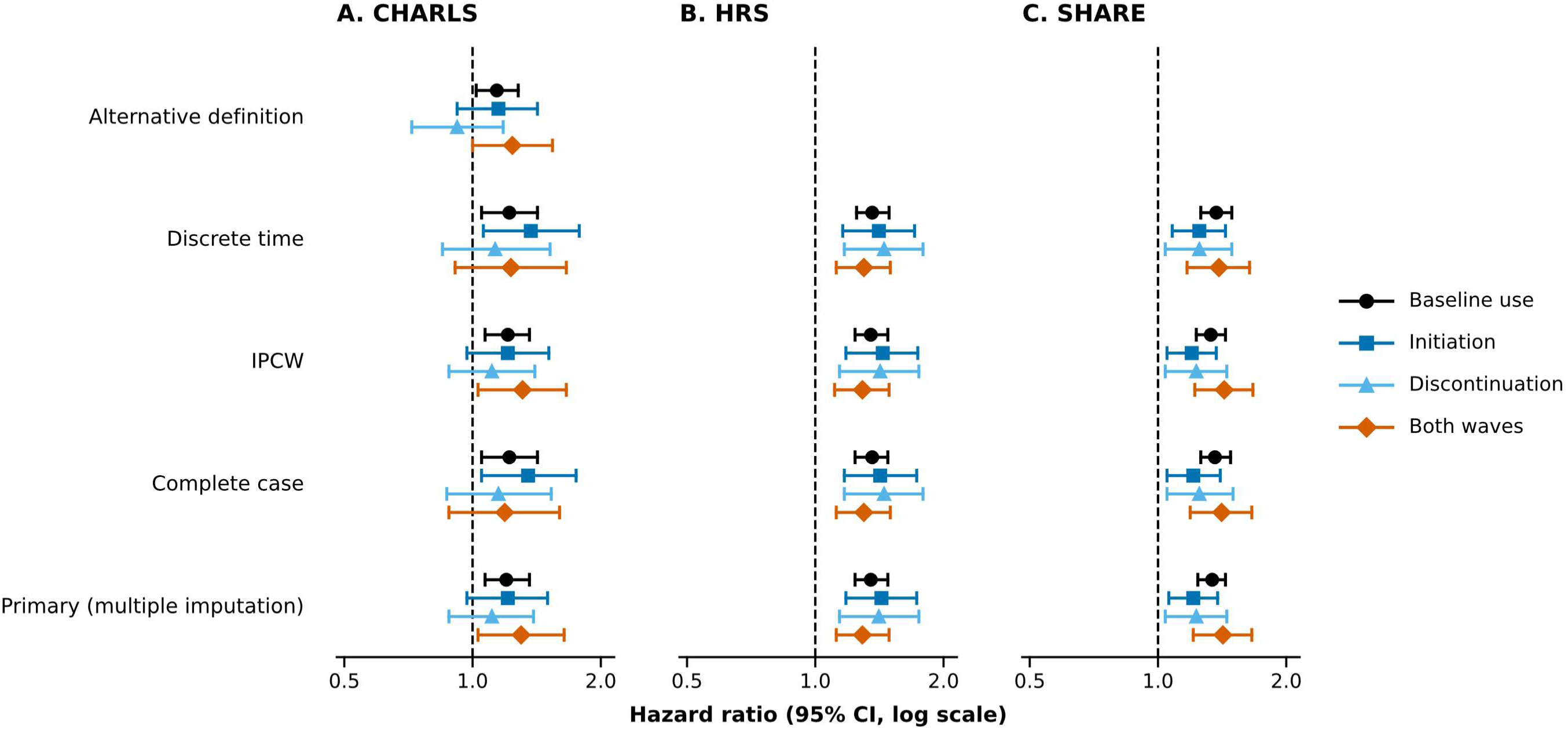
Sensitivity estimates across alternative analytic specifications. Hazard ratios (Model 4) with 95% confidence intervals are shown on a logarithmic scale for baseline gastrointestinal medication use and the three two-wave patterns (initiation, discontinuation and use reported at both waves) in (A) CHARLS, (B) HRS and (C) SHARE. Rows present the primary multiple-imputation model, complete-case analysis, stabilised inverse-probability weighting (IPCW), an observed-visit discrete-time complementary log-log model, and (for CHARLS) the broader alternative medication definition.

**Figure S3.**
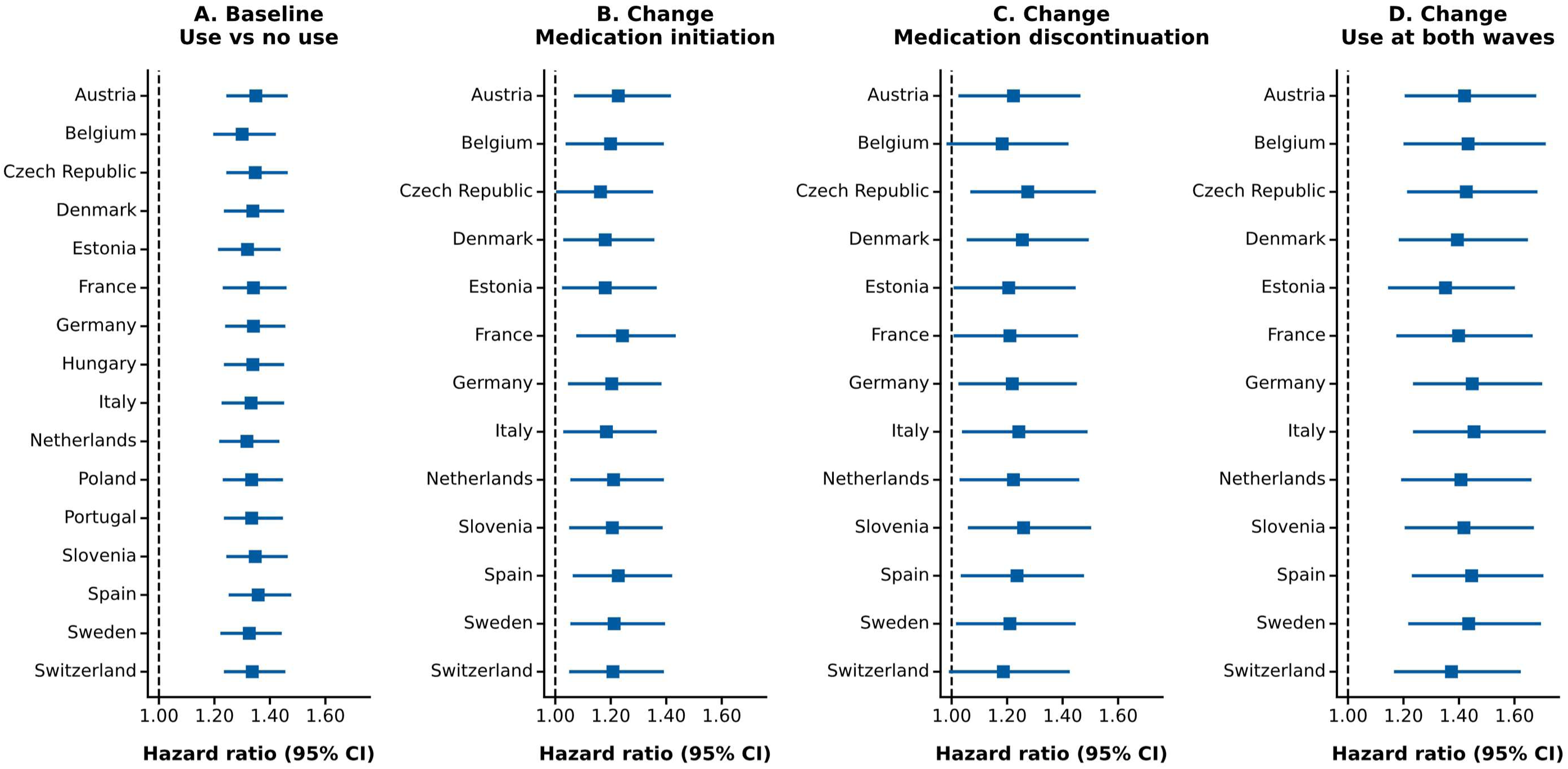
SHARE leave-one-country-out stability analysis. Each estimate omits one SHARE country and refits the primary Model 4; the display evaluates whether a single country dominates the cohort-specific result.

**Figure S4.**
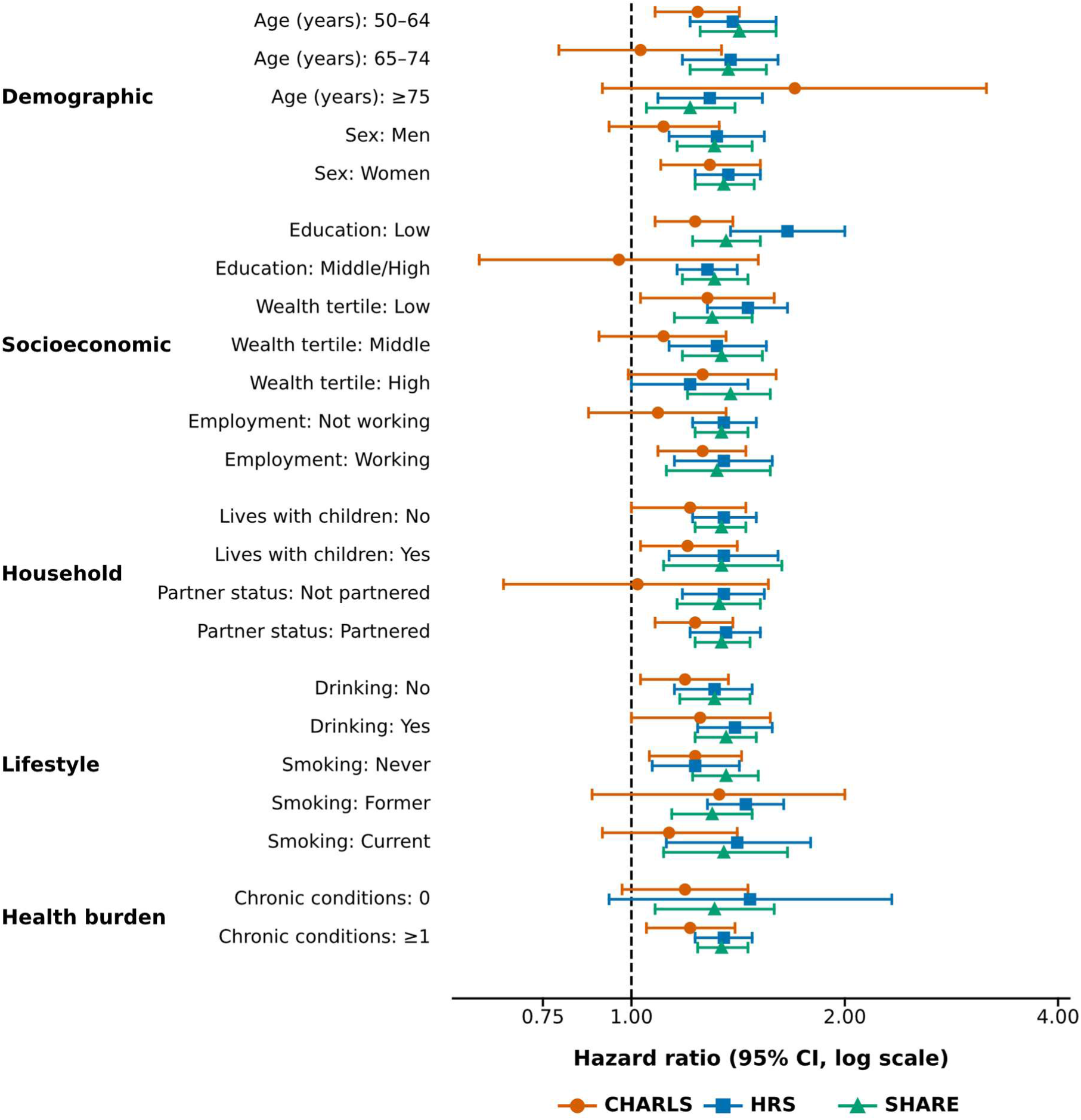
Baseline gastrointestinal medication-use subgroup analysis. Points show cohort-specific Model 4 hazard ratios with 95% confidence intervals within prespecified baseline subgroups, offset vertically within each stratum. Tests for effect modification (*P* for interaction) are reported in Table S13.

**Figure S5.**
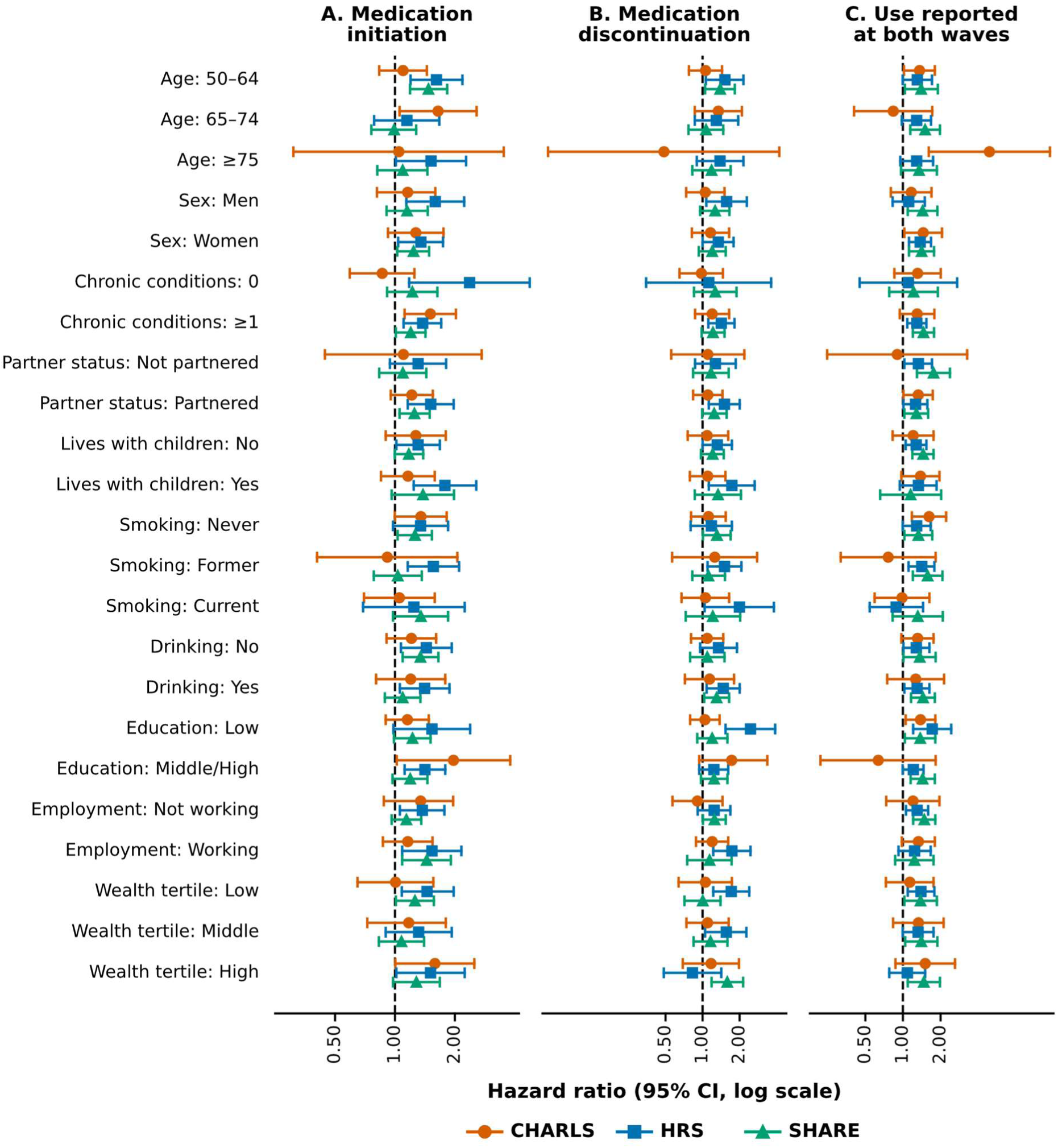
Two-wave medication-use patterns and incident depressive symptoms, by subgroup. Points show stratum-specific hazard ratios (Model 4) with 95% confidence intervals for CHARLS (circles), HRS (squares) and SHARE (triangles); panels show (A) medication initiation, (B) medication discontinuation and (C) use reported at both waves (versus continued non-use). Estimates are displayed graphically because stratum-specific groups were small, particularly in CHARLS.

**Figure S6.**
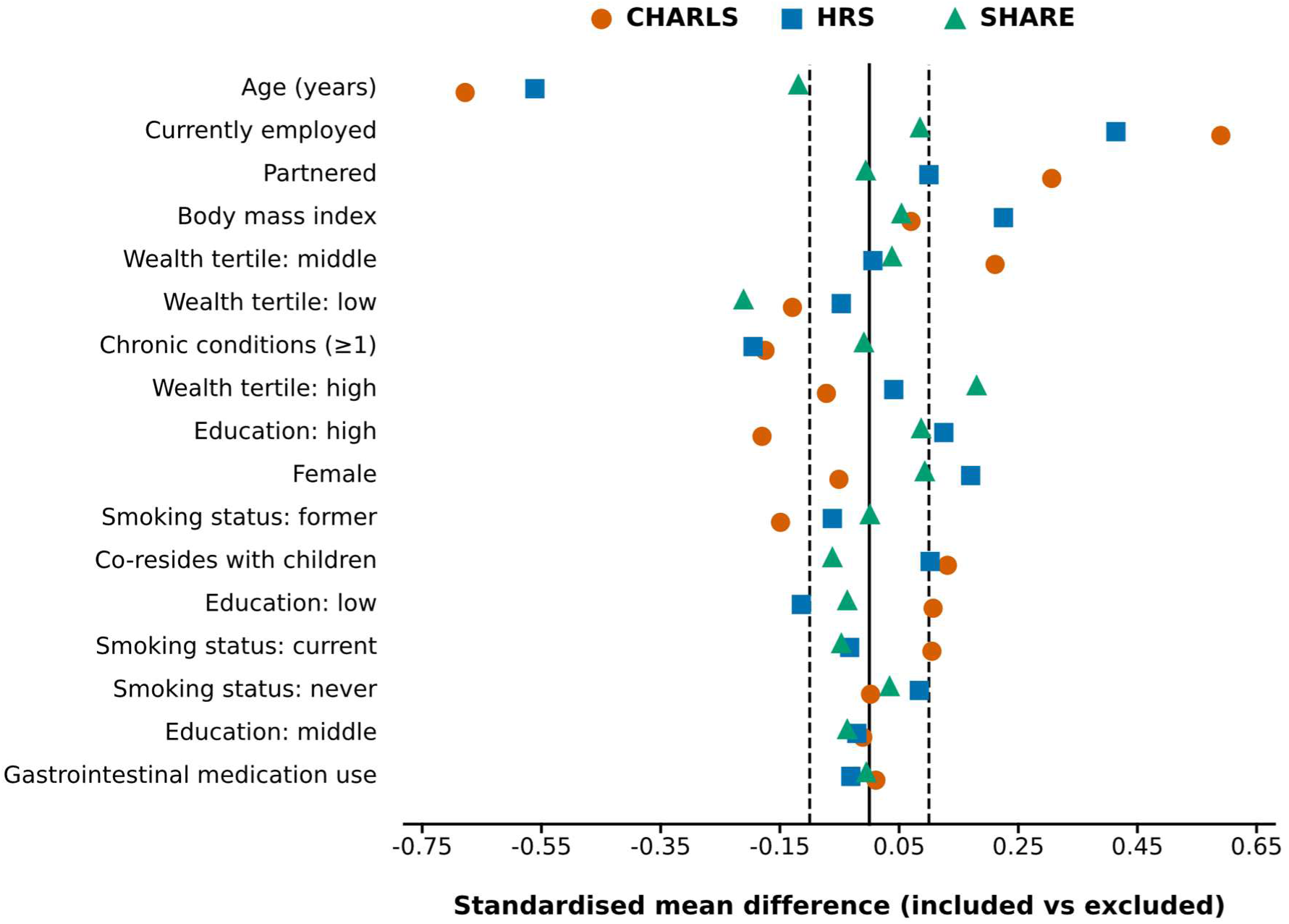
Standardised mean differences in baseline characteristics between participants included in and excluded from the analytic samples, by cohort. Dashed lines indicate an absolute standardised mean difference of 0.10.

### Additional file 2

**STROBE Statement—checklist of items that should be included in reports of cohort studies**

*Manuscript: Gastrointestinal medication use patterns and incident depressive symptoms among adults aged 50 years or older: a prospective multicohort study*

This checklist follows the Strengthening the Reporting of Observational Studies in Epidemiology (STROBE) statement for cohort studies (von Elm et al., 2007). Each item is mapped to the relevant section, table, or figure in the manuscript. Section names and table/figure numbers are used as stable locators; page numbers are omitted because pagination changes between Word and journal typesetting.

| Section | Item No. | Recommendation | Reported on section/table/figure |
| --- | --- | --- | --- |
| **Title and abstract** | 1 (a) | Indicate the study's design with a commonly used term in the title or the abstract. | Title includes the design term “a prospective multicohort study”; the design is restated in the Abstract. |
| **Title and abstract** | 1 (b) | Provide in the abstract an informative and balanced summary of what was done and what was found. | Structured Abstract—Background, Methods, Results, and Conclusions, reporting sample sizes, cohort-specific and pooled hazard ratios, negative/positive controls, and competing-risk analyses. |
| **Introduction** | 2 | Explain the scientific background and rationale for the investigation being reported. | Background, paragraphs 1–3: burden of late-life depression, gastrointestinal illness and medication use, and the three identified evidence gaps. |
| **Introduction** | 3 | State specific objectives, including any prespecified hypotheses. | Background, final paragraph: three prespecified aims (1)–(3), addressing baseline and two-wave associations, specificity via controls, and robustness. |
| **Methods** | 4 | Present key elements of study design early in the paper. | Methods, “Study design and participants”: parallel prospective longitudinal analyses of three harmonised ageing cohorts, analysed separately then pooled. |
| **Methods** | 5 | Describe the setting, locations, and relevant dates, including periods of recruitment, exposure, follow-up, and data collection. | “Study design and participants”: China (CHARLS, 2011–2020), United States (HRS, 2012–2020), and 16 European countries (SHARE, 2010/11–2021/22); baseline and second-round medication waves, follow-up waves; Tables S1 and S21. |
| **Methods** | 6 (a) | Cohort study: Give the eligibility criteria, and the sources and methods of selection of participants. Describe methods of follow-up. | “Study design and participants”: age $\geq 50$ years, baseline medication and symptom-score availability, below-threshold baseline score, and $\geq 1$ follow-up assessment; two-wave criteria and analytic cohort construction. Follow-up in “Depressive-symptom outcome and follow-up”. Flow: Fig. 1; Fig. S1. |
| **Methods** | 6 (b) | Cohort study: For matched studies, give matching criteria and number of exposed and unexposed. | Not applicable—no matching was used; exposed and unexposed groups were defined by reported medication status within each cohort. |
| **Methods** | 7 | Clearly define all outcomes, exposures, predictors, potential confounders, and effect modifiers. Give diagnostic criteria, if applicable. | “Medication exposure”; “Depressive-symptom outcome and follow-up” (CES-D and EURO-D thresholds $\geq 10/\geq 3/\geq 4$); “Covariates”; harmonised definitions in Table S1. |
| **Methods** | 8 | For each variable of interest, give sources of data and details of methods of assessment (measurement). Describe comparability of assessment methods if there is more than one group. | “Medication exposure”: cohort-specific questionnaire wording for CHARLS, HRS, and SHARE; outcome instruments and cross-cohort comparability in “Depressive-symptom outcome and follow-up”; Table S1. |
| **Methods** | 9 | Describe any efforts to address potential sources of bias. | “Subgroup and sensitivity analyses”; negative- and positive-control exposures; “Competing risk”; “Missing data”; standardised-difference comparison of included vs eligible participants (Fig. S6). |
| **Methods** | 10 | Explain how the study size was arrived at. | “Study design and participants”: all eligible participants meeting prespecified inclusion criteria in the three national cohorts were included (50,467 in the baseline analysis; 32,686 in the two-wave analysis); cohort-specific derivation is shown in Fig. 1 and Fig. S1. |
| **Methods** | 11 | Explain how quantitative variables were handled in the analyses. If applicable, describe which groupings were chosen, and why. | “Covariates”: age and BMI modelled continuously; educational attainment in three harmonised tiers; household wealth in within-cohort tertiles; chronic-condition burden dichotomised (0 vs $\geq 1$). Natural cubic spline age and categorical chronic-condition counts (0, 1, 2, $\geq 3$) evaluated in “Primary Cox proportional-hazards analyses” (Tables S17 and S20). |
| **Methods** | 12 (a) | Describe all statistical methods, including those used to control for confounding. | “Statistical analysis” and “Primary Cox proportional-hazards analyses”: cohort-specific Cox models, cluster-robust standard errors, SHARE country fixed effects, cumulative Models 1–4; “Cross-cohort synthesis”. |
| **Methods** | 12 (b) | Describe any methods used to examine subgroups and interactions. | “Subgroup and sensitivity analyses”: exposure-by-subgroup interaction terms; Results interaction tests; Table S13 and Figs. S4–S5. |
| **Methods** | 12 (c) | Explain how missing data were addressed. | “Missing data”: multiple imputation by chained equations (20 datasets; estimates combined by Rubin's rules); descriptive tables use available denominators. |
| **Methods** | 12 (d) | Cohort study: If applicable, explain how loss to follow-up was addressed. | “Competing risk”, inverse-probability-of-censoring-weighted (IPCW) Cox models, and standardised-difference selection analysis (Fig. S6); Results. |
| **Methods** | 12 (e) | Describe any sensitivity analyses. | “Subgroup and sensitivity analyses”: complete-case, IPCW, complementary log-log, rolling person-period, broader CHARLS exposure, digestive-disease cross-classification, leave-one-country-out, and E-values; Tables S8–S11, S16–S20 and Figs. S2–S3. |
| Results | 13 (a) | Report the numbers of individuals at each stage of the study—e.g., numbers potentially eligible, examined for eligibility, confirmed eligible, included in the study, completing follow-up, and analysed. | “Participants and gastrointestinal medication-use patterns”: 6,452 / 12,938 / 31,077 (baseline) and 4,087 / 9,801 / 18,798 (two-wave); Fig. 1; Fig. S1. |
| Results | 13 (b) | Give reasons for non-participation at each stage. | Eligibility rules in Methods; cohort-specific exclusion reasons and counts in Fig. S1 (missing medication/symptom data, no valid follow-up, and deaths before the first follow-up). |
| Results | 13 (c) | Consider use of a flow diagram. | Fig. 1 (analysis timelines and sample sizes); detailed cohort-specific flow in Fig. S1. |
| Results | 14 (a) | Give characteristics of study participants (e.g., demographic, clinical, social) and information on exposures and potential confounders. | Table 1, panels A–B; exposure-stratified characteristics in Tables S2–S7. |
| Results | 14 (b) | Indicate the number of participants with missing data for each variable of interest. | “Missing data”; missingness rates for all covariates in the baseline analytic sample in Table S22; denominators in Table 1 and Tables S2–S7; missing covariates multiply imputed. |
| Results | 14 (c) | Cohort study: Summarise follow-up time (e.g., average and total amount). | Table 1: person-years 39,725 / 75,700 / 209,762 and median follow-up 9, 8, and 11 years for CHARLS, HRS, and SHARE; Results. |
| Results | 15 | Cohort study: Report numbers of outcome events or summary measures over time. | Results and Table 1: 16,896 incident events (baseline; 8,069 two-wave) and incidence rates per 1,000 person-years (72.6, 44.4, 50.8); hazard ratios over time in Table 2. |
| Results | 16 (a) | Give unadjusted and, if applicable, confounder-adjusted estimates and their precision (e.g., 95% confidence interval). Make clear which confounders were adjusted for and why they were included. | “Primary associations”: unadjusted Model 1 through fully adjusted Model 4 hazard ratios with 95% CIs; confounders specified in Methods; Table 2; Fig. 2; Table S12. |
| Results | 16 (b) | Report category boundaries when continuous variables were categorised. | “Covariates”; boundaries in Table 1, Table S1 (education tiers, within-cohort wealth tertiles, symptom thresholds), and subgroup tables (Table S13). |
| Results | 16 (c) | If relevant, consider translating estimates of relative risk into absolute risk for a meaningful time period. | Table 1 reports person-years of follow-up, incident event counts, and unadjusted incidence rates per 1,000 person-years in each cohort alongside relative hazard ratios in Table 2. |
| Results | 17 | Report other analyses done—e.g., analyses of subgroups and interactions, and sensitivity analyses. | “Sensitivity and secondary analyses”, “Negative- and positive-control exposures”, and “Competing risk” results; Table 3; Fig. 3; Tables S8–S20 and Figs. S2–S6. |
| Discussion | 18 | Summarise key results with reference to study objectives. | Discussion, first paragraph; Conclusions. |
| Discussion | 19 | Discuss limitations of the study, taking into account sources of potential bias or imprecision. Discuss both direction and magnitude of any potential bias. | Discussion, “Several limitations...”: evaluation of potential biases and methodological considerations, including non-differential exposure misclassification and residual confounding. |
| **Discussion** | 20 | Give a cautious overall interpretation of results considering objectives, limitations, multiplicity of analyses, results from similar studies, and other relevant evidence. | Discussion: balanced interpretation integrating biological mechanisms, underlying digestive morbidity, negative/positive controls, and clinical implications for routine affective screening during medication reviews; Conclusions. |
| **Discussion** | 21 | Discuss the generalisability (external validity) of the study results. | Discussion: cross-national generalisability across China, the United States, and 16 European countries spanning diverse healthcare systems, welfare regimes, and validated symptom scales; 95% prediction intervals reported in Results and Table S12; Conclusions. |
| **Other information** | 22 | Give the source of funding and the role of the funders for the present study and, if applicable, for the original study on which the present article is based. | Funding; Availability of data and materials; Authors' contributions, including the guarantor's role. |

Reference: von Elm E, Altman DG, Egger M, Pocock SJ, Gøtzsche PC, Vandenbroucke JP. The Strengthening the Reporting of Observational Studies in Epidemiology (STROBE) Statement: guidelines for reporting observational studies. *Ann Intern Med.* 2007;147:573–7. https://doi.org/10.7326/0003-4819-147-8-200710160-00010.

## Notes

### Competing Interest Statement

The authors have declared no competing interest.

### Author Declarations

The Institutional Review Board of Peking University (approval number IRB00001052-11015) gave ethical approval for the China Health and Retirement Longitudinal Study (CHARLS). The Health Sciences and Behavioral Sciences Institutional Review Board of the University of Michigan gave ethical approval for the Health and Retirement Study (HRS). The Ethics Committee of the University of Mannheim and the Ethics Council of the Max Planck Society for the Advancement of Science gave ethical approval for the Survey of Health, Ageing and Retirement in Europe (SHARE). All participants provided informed consent in the original cohort studies, and this work analysed de-identified data obtained via registered researcher access.

